# Quantitative Gradient Echo MRI Identifies Dark Matter as a New Imaging Biomarker of Neurodegeneration that Precedes Tissue Atrophy in Early Alzheimer Disease

**DOI:** 10.1101/2021.04.27.21256098

**Authors:** Satya V.V.N. Kothapalli, Tammie L. Benzinger, Andrew. J. Aschenbrenner, Richard. J. Perrin, Charles. F. Hildebolt, Manu. S. Goyal, Anne. M. Fagan, Marcus. E. Raichle, John. C. Morris, Dmitriy. A. Yablonskiy

## Abstract

**Background:** Currently, brain tissue atrophy serves as in vivo MRI biomarker of neurodegeneration in Alzheimer Disease (AD). However, postmortem histopathological studies show that neuronal loss in AD exceeds volumetric loss of tissue and that loss of memory in AD begins when neurons and synapses are lost. Therefore, in vivo detection of neuronal loss prior to detectable atrophy in MRI is essential for early AD diagnosis.

**Objective:** To apply a recently developed quantitative Gradient Recalled Echo (qGRE) MRI technique for in vivo evaluation of neuronal loss in human hippocampus.

**Methods:** Seventy participants were recruited from the Knight Alzheimer Disease Research Center, representing three groups: Healthy controls [Clinical Dementia Rating® (CDR®)=0, amyloid β (Aβ)-negative), n=34]; Preclinical AD (CDR=0, Aβ-positive, n=19); and mild AD (CDR=0.5 or 1, Aβ-positive, n=17).

**Results:** In hippocampal tissue, qGRE identified two types of regions: one, practically devoid of neurons, we designate as “Dark Matter”, the other, with relatively preserved neurons - “Viable Tissue”. Data showed a greater loss of neurons than defined by atrophy in the mild AD group compared with the healthy control group - neuronal loss ranged between 31% and 43% while volume loss ranged only between 10% and 19%. The concept of Dark Matter was confirmed with histopathological study of one participant who underwent in vivo qGRE 14 months prior to expiration.

**Conclusion:** in vivo qGRE method identifies neuronal loss that is associated with impaired AD-related cognition but is not recognized by MRI measurements of tissue atrophy, therefore providing new biomarkers for early AD detection.

## Introduction

Alzheimer disease (AD) is a progressive neurodegenerative disorder with typical clinical symptoms of memory loss, global cognitive decline, and behavioral changes, which eventually impact daily living activities [1]. It is well-known that pathological changes in the human brain begin decades before the appearance of clinical AD symptoms [2-6]. Magnetic resonance imaging (MRI) studies indicate that hippocampal atrophy (loss of tissue) is one of the earliest indications of AD pathology that can be detected through morphological studies [7-10]. The hippocampus is a complex structure that consists of several subregions that include dentate gyrus (DG), cornu ammonis (CA1, CA2, CA3, and CA4), subiculum and molecular layer (ML) [11]. Importantly, neurofibrillary tangles (NFT), a hallmark of AD, typically demonstrate a characteristic pattern of evolution across hippocampal subfields--first appearing in the CA1 and then spreading to other subfields [12, 13]. The hippocampal subfields have different functions and network connections [11, 14, 15] and show different vulnerabilities to neuropathologic insults (e.g., CA4 and CA1 to hypoxic/ischemic injury and excitotoxicity). Advances in MRI acquisition and image processing techniques allow segmentation of these sub-regions for investigating their volumetric and morphometric properties in relation to various neurodegenerative diseases [16]. Reports have shown that different hippocampal subfields exhibit differential vulnerability to various neurodegenerative disease pathologies, and variable involvement at different stages of these diseases [17, 18]. In AD, however, morphometric alternations have been detected in multiple hippocampal subfields [15, 19-21] and were associated with mild cognitive impairment [22, 23].

While volumetric MRI-based measurement of atrophy is usually assumed to serve as an in vivo biomarker of neuronal loss [24], histopathological studies demonstrate that neuronal loss in the hippocampus actually exceeds loss of tissue volume [25]. This result is consistent with the finding of a widespread reduction of synapses that were more extensive than decreases in gray matter volume [26]. Importantly, symptoms of Alzheimer disease appear after sufficient neuronal [25, 27] and synaptic [28, 29] losses have occurred. Additionally, studies demonstrate that the extent of neuronal loss varies across hippocampal subfields, e.g., severe neuronal loss is observed in CA1 with less severe loss in subiculum regions [30]. Hence, in vivo assessment of neuronal loss in hippocampal subfields may serve as a more effective biomarker for disease progression than will measures of atrophy.

Our approach to assessing neuronal damage in vivo is based on (a) the quantitative Gradient Recalled Echo (qGRE) MRI technique [31] and (b) genetically-informed quantitative relationships between qGRE metrics and major components of brain tissue cellular structure (neurons/neurites and glia) [32]. The qGRE technique provides quantitative, non-invasive, in vivo, high-resolution 3D measurements of several brain-tissue-specific relaxation properties (qGRE metrics) of the gradient recalled echo (GRE) MRI signal that depend on brain cellular structure and functioning. Specifically, qGRE separates the GRE MRI signal decay R2* (=1/T2*) into its two main components, R2t* (t stands for tissue) and R2′. The R2t* relaxation parameter depends solely on the cellular and subcellular microstructure [32], whereas R2′ relates to the GRE signal loss due to the presence of paramagnetic deoxyhemoglobin in venous blood and adjacent to it part of the capillary bed (BOLD effect) [33, 34]. The basis of R2* disentanglement into R2t* and R2’ is grounded in our previously developed theoretical model of BOLD signal [34] validated in phantom [35] and animal [36] studies. By introducing a concept of cells as an endogenous contrast agent, a quantitative relationship between the R2t* qGRE metric and Neuronal Density Index (NDI) – a parameter that represents a proxy for the neuronal density – was established in Ref. [32] (see further details below in the Methods section).

In this study, we demonstrate that the qGRE R2t* metric identifies two different types of tissues in the hippocampal subfields of people with preclinical and mild AD dementia: one type – tissue with markedly lower neuronal content (that we term “Dark Matter” as it appears dark on R2t* images), and another type – tissue with a relatively preserved concentration of neurons (that we term “Viable Tissue”). We demonstrate that the qGRE-measured volumes of Viable Tissue and Dark Matter account for more significant differentiation between healthy participants and people with preclinical and mild AD than the total volume measurements by morphometric MRI. Likewise, the volumes of Viable Tissue and Dark Matter have stronger association with memory scores as compared with total volume. Finally, our preliminary data show that premortem in vivo R2t*-based measurement of neuronal content in hippocampal subfields correspond to postmortem neuronal counts in an individual investigated by histopathology.

## Results

Detailed procedures of generating qGRE-based metrics of Dark Matter, Viable Tissue and Neuronal Density Index are provided in the Methods section.

### AD-related neurodegeneration (neuronal damage) affects all hippocampal subfields

The group comparisons of Dark Matter fraction, total volume, and Viable Tissue volume measurements between HC, PC, and mild AD groups in the hippocampal subfields are presented in **Figure 1**. We found no significant differences (p>0.05) between the HC and PC groups in total volume, which is in good agreement with previous histological [25] and longitudinal MRI [37] studies. Consistent with previous reports [21, 38, 39], we also found decreased volume in all hippocampal subfields in the mild AD group compared with the PC and HC groups though in some subfields (e.g. CA1) these differences did not reach the significance threshold (most likely due to the small sample sizes). This result is consistent with both 3T [38] and 7T MRI studies of hippocampal subfield volumes [40]. The significant volume differences between the mild AD and HC groups in our data are seen in all subfields except in CA1 and CA2/3 regions. Further, group differences (p<0.05) between mild AD and PC are seen in all subfields except in CA1. Compared with total tissue volume, Viable Tissue, and Dark Matter volumes show stronger group separation in most hippocampal subfields (**Figure 1**). Unlike total volume measurements, the Viable Tissue volumes in all hippocampal subfields in the mild AD group differ from those of the HC group and PC group (p<0.05). The Dark Matter fraction exhibits group differences between mild AD and HC (p<0.01) in all subfields and exhibits differences between PC and mild AD (p<0.05) in CA1, CA2/3, CA4, GC-DG, and ML regions. Importantly, statistically significant group differences in Dark Matter fraction were found between HC and PC (p<0.05) in the parasubiculum, subiculum, CA1, and ML regions.

**Figure 1.**
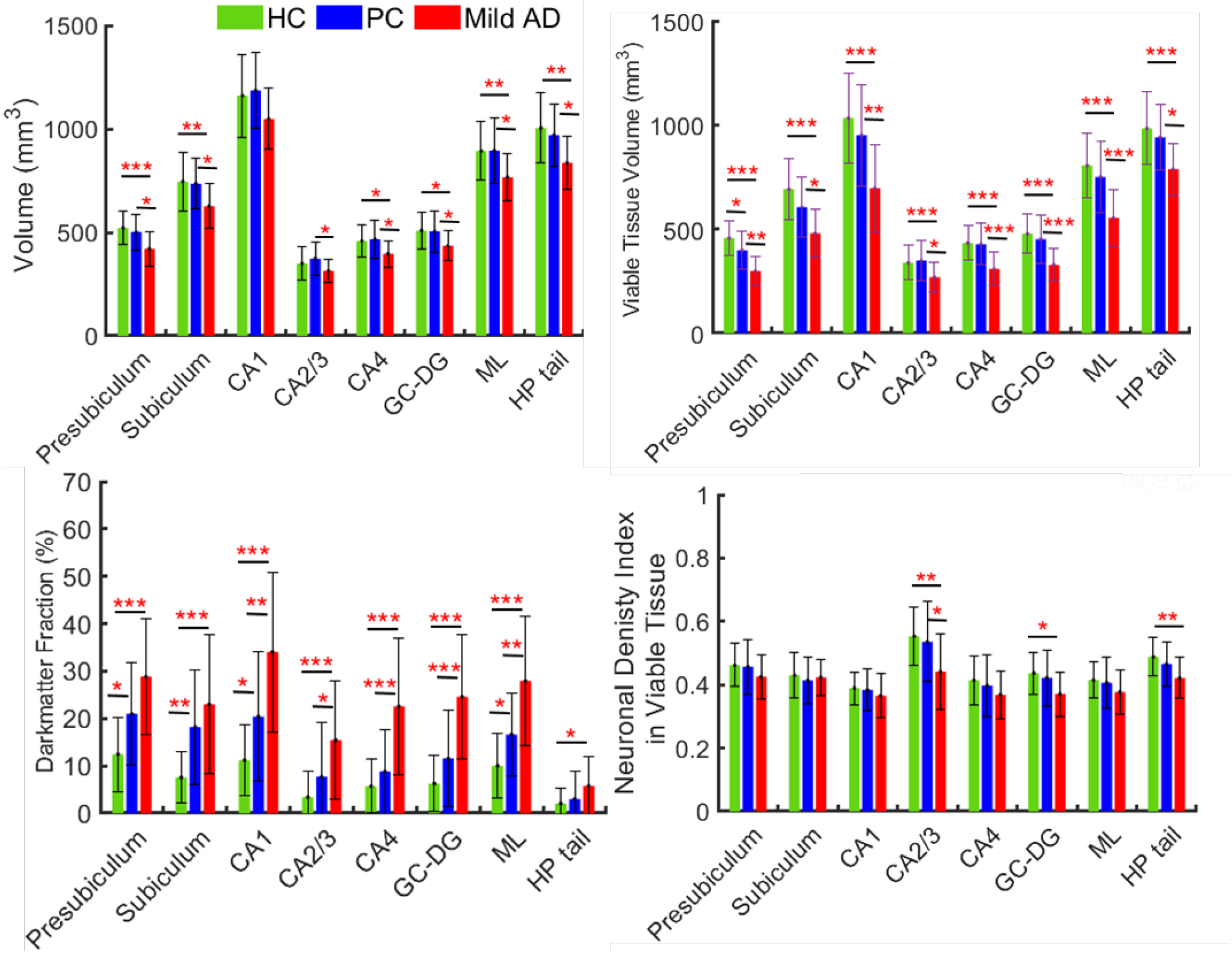
Total volume, Viable Tissue volume, Dark Matter fraction, and Neuronal Density Index of the viable tissue in hippocampal subfields. Bars represent mean group values and whiskers show standard deviations. Green bars represent the HC group (n=34), blue bars represent PC group (n=19), and red bars represent mild AD (n=17) group. Herein, the “**\***” corresponds to p<0.05, “**\*\***” to p<0.01, “**\*\*\***” to p<0.001.

The last panel in **Figure 1** represents measurements of neuronal density in the Viable Tissue of all hippocampal subfields. It is evident that mean value of NDI of the Viable Tissue in mild AD is decreased as compared with HC or PC. However, the one-way ANOVA multi comparison identified significant differences between HC and mild AD observed only in specific subfields, such as CA2/3, GC-DG, and HP tail regions, and significant group differences between PC and mild AD was observed only in CA2/3 region. This finding remarkably indicates that neurons are only marginally decreased in the Viable Tissue while major neuronal loss is related to the presence of Dark Matter. The lower mean NDI in the mild AD group might be attributed to increased microglia cell activity in the Viable Tissue in mild AD compared with HC or PC tissue.

### Neuronal loss exceeds volumetric loss of tissue in hippocampal subfields

Data in **Figure 2** shows loss of neurons and tissue volume in the mild AD group relative to the HC group. The mean volume loss fraction of the mild AD group is about 19% in presubiculum, 16% in subiculum, 10% in CA1, 10% in CA2/3, 14% in CA4, 15% in GC-DG, 14% ML, and 17% in HP tail with respect to the HC group. Previous studies have reported that the overall hippocampal volume was lower by 15-30% in mild AD compared with HC [41]. In our findings, a volume reduction was about 10-19% in the mild AD group compared with the HC group, which is consistent with previous findings. The mean neuronal loss fraction in the mild AD group is about 40% in presubiculum, 32% in subiculum, 37% in CA1, 37% in CA2/3, 38% in CA4, 43% in GC-DG, 38% ML, and 31% in HP tail. These data show significantly higher neuronal loss as compared with volume loss in the mild AD group. This result is consistent with previous findings [25] that reported about 46% of neuronal loss and 29% of volumetric loss (about 1.6 times lower than neuronal loss) in the CA1 region in the AD postmortem brains compared with cognitively normal brains. The presence of Dark Matter (i.e. brain tissue with significantly lower concentration of neurons with their processes, hence less restrictions for water diffusion) can also explain previously reported measurements of increased Apparent Diffusion Coefficient in hippocampus of people with mild cognitive impairment [42].

**Figure 2.**
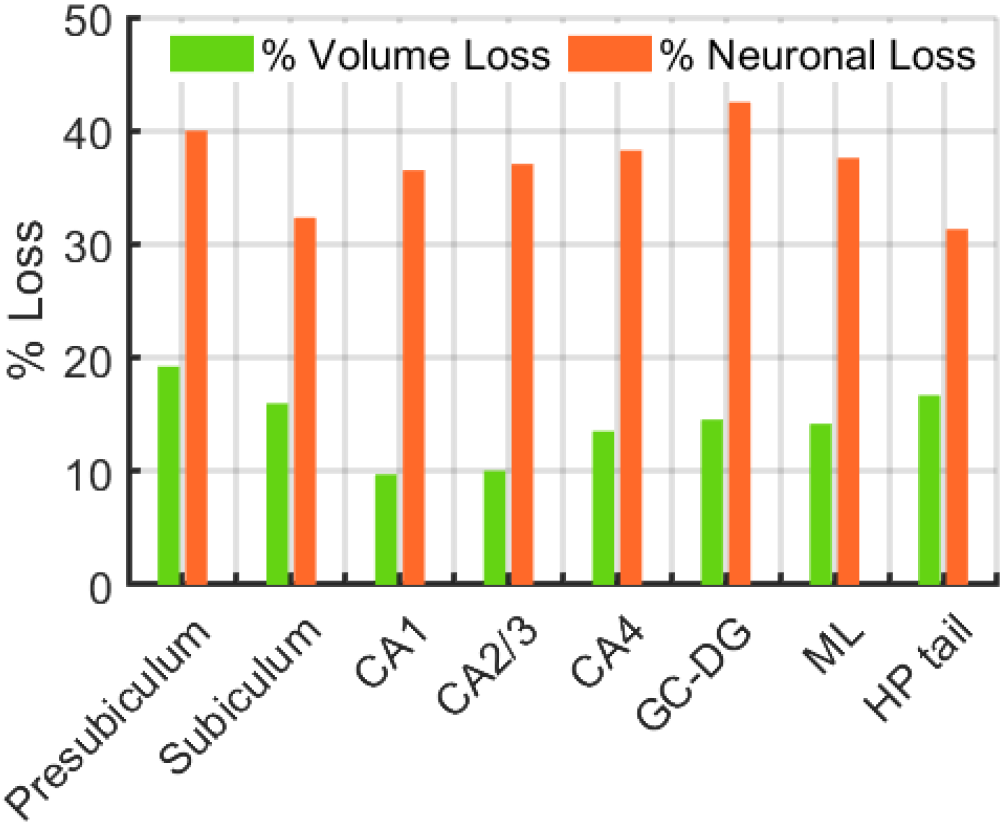
Percent of volume and neuronal losses in mild AD group in hippocampal subfields evaluated with respect to the group mean values of HC group.

### Dark Matter fraction and the Viable Tissue volume exhibit stronger associations with neurocognitive scores than volumetric measurements

The outcomes of linear regression analyses between the global cognitive composite and the Free and Cued Selective Reminding Test, (FcSRT, as a measure of episodic memory) and imaging scores (qGRE biomarkers of Viable Tissue volume and Dark Matter fraction, along with the Total Volume measurements in global hippocampus) are presented in **Figure 3**. Outcomes from the individual cognitive tests are presented in Supplemental Figure S1. The Dark Matter fraction is negatively associated with episodic memory (R=-0.47; p=0.0001), meaning that higher Dark Matter fraction correlates with worse memory performance, while the Total Volume exhibits a positive but much weaker association (R=0.23; p=0.07), i.e., lower the volume lower the episodic memory scores. A similar trend is seen for the global cognitive composite, though in this case, the Dark Matter fraction and the Total Volume shows similar correlations, while the strongest association is seen with the Viable Tissue Volume (R=0.53, p < 0.0001).

**Figure 3.**
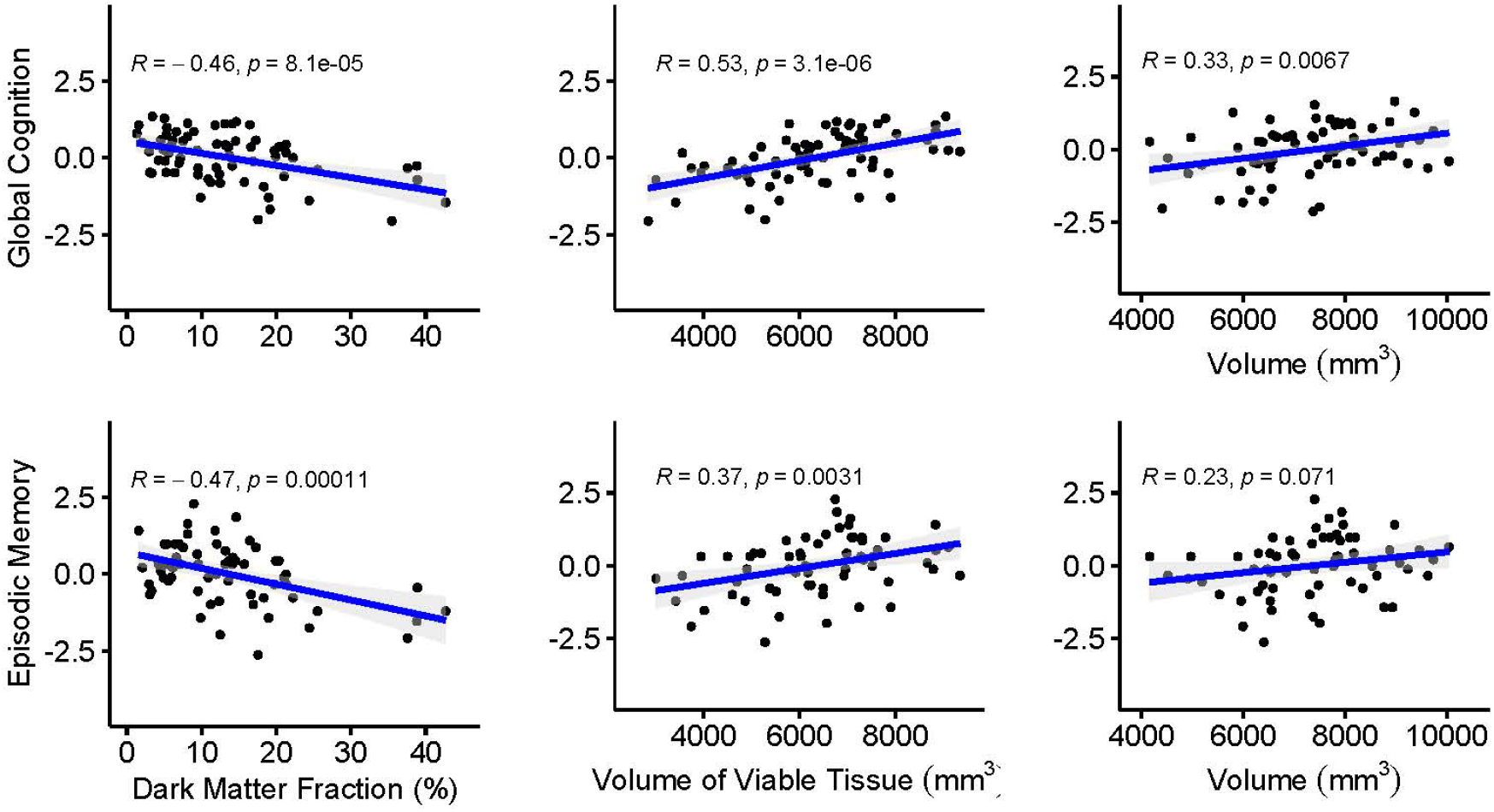
Correlation of two cognitive scores: z-scores of Global Cognition and Episodic Memory (represented by the Global Memory Composite and the Free and Cued Selective Reminding Test, respectively) vs. fraction of Dark Matter, volume of Viable Tissue, and Total Volume of the hippocampus. Each point represents an individual participant. All individual cognitive data are presented in Supplemental Figure S1.

Our results for the Total Volume measurements indicate that smaller hippocampal volumes are associated with worse cognitive performance, consistent with the previous studies [43] [44]. Plots in **Figure 3** and Supplemental Figure S1 show a positive association with FcSRT, global cognition, letter number sequencing, category fluency and negative associations (as expected) with the Trail Making A and B tests.

### Volumes of Hippocampal Dark Matter and Viable Tissue provide new biomarkers for classification of HC, PC, and mild AD groups

In this section, we present the results of our second set of data analyses that are based on global hippocampal measures and use these biomarkers to demonstrate that the new qGRE metrics (Dark Matter and Viable Tissue volumes) represent clinically important differences between groups as compared with differences based on hippocampal atrophy (reduction of hippocampal volume). Results based on the values of the individual biomarkers are shown in **Figure 4**. The box plots illustrate the clinical importance of differences among the values for the HC, PC and mild AD groups in the hippocampus. These differences among groups were assessed with a generalized linear model with a normal distribution and an identity link function. As part of this analysis, to assess differences between groups, contrast analyses were performed. Although the use of a generalized linear model does not require the assumptions of data normality or equality of variances, for each model a studentized deviance residual plot was created and examined to detect trends that were not captured by the model. For these analyses, the data distributions in the studentized deviance residual plots were judged acceptable. To partially control for these multiple assessments, alpha was set at 0.01.

**Figure 4.**
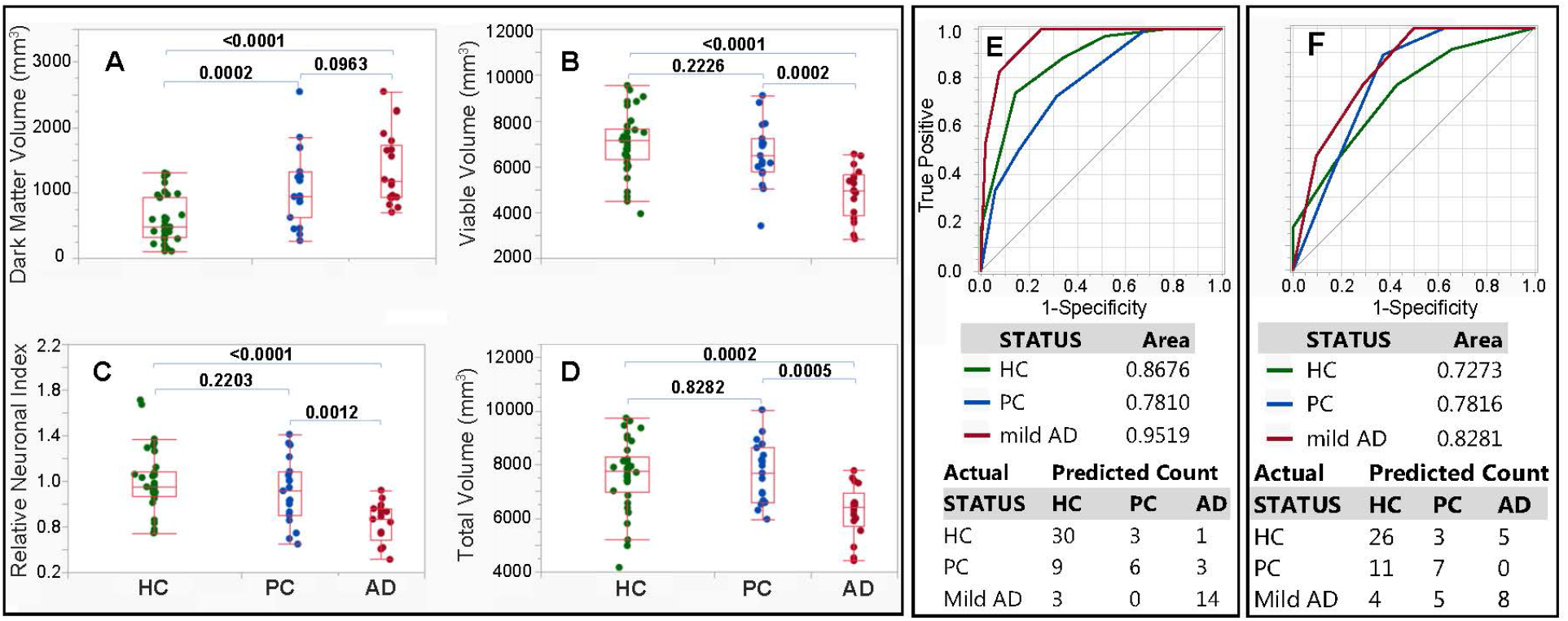
Group differences (A-D) and ROC classifications (E, F) based on qGRE metrics and volumetric measurements in the hippocampus. (**A**) Dark Matter volume (mm^3^), (**B**) Viable Tissue volume (mm^3^), (**C**) Relative Neuronal Index (total number of neurons normalized by the mean value of the total number of neurons in the HC group), and (D) Total Volume (mm^3^). The middle lines of the box plots represent median, ends of the boxes are the 25th and 75th quantiles (quartiles), and interquartile range is the difference between the quartiles. The lines (whiskers) extend from the boxes to the outermost points that fall within the distance computed as 1.5 (interquartile range). All generalized linear models for A, B, C, and D indicated differences among the HC, PC, and AD groups (p ≤ 0.0004) and the p values that resulted from assessing differences between groups are indicated above the horizontal connectors between groups. (**E**) Result of a classification-tree that was produced using global hippocampal Dark Matter volume and Viable Tissue volume variables as predictors. (**F**) Result of a classification-tree that was produced using Total Hippocampal Volume as predictor. Receiver operating characteristic (ROC) curves, areas under the curves (AUCs), and a confusion matrix are presented. The confusion matrix presents the numbers of correct and incorrect classifications. ROC analysis is performed by constructing a graph of true and false positive rates (sensitivity and 1 minus specificity, respectively) for a series of cutoff points for a test (in our case, MRI metrics). The ROC curve yields a measure of diagnostic accuracy, independent of the decision criterion. It characterizes the inherent accuracy of the technique. The AUC value represents the probability that a randomly chosen abnormal case is (correctly) rated or ranked with greater suspicion than a randomly chosen normal case.

The findings demonstrate that while total hippocampal volume results in statistically significant differences between the mild AD group and the HC and PC groups, it does not result in a statistically significant difference between the PC and HC groups. At the same time, the Dark Matter volume is significantly elevated in PC group as compared with the HC group and may, thus, serve as a biomarker for early AD pathology. Note that, even after excluding the outlier with unusually high Dark Matter Volume (2542 mm^3^ – higher than any other participant in our study, including participants with AD) from the PC group (**Figure 4**A), the difference between the HC and PC groups remains highly significant (p=0.0008), and the p value for the difference between the PC and mild AD groups decreases (p=0.0218 vs. 0.0963 in **Figure 4**A). Though, the outlier participant is cognitively normal, her very high Dark Matter volume is consistent with elevated tauopathy standard uptake value ratio (1.77) and the lower episodic memory score, which is close to the mean cognitive score of the mild AD group (mean FcSRT score of mild AD group was about 16.5, and the FcSRT score for this participant was 16). Presumably, this participant might progress to symptomatic AD very soon, or might have unknown factors conferring cognitive resilience. This outlier was further excluded from classification tree analysis.

Results for the neuronal content show similar values in the HC and PC groups but an impressively lower number of neurons in the mild AD participants. These assessments were performed to estimate population parameters.

In addition, models were used to calculate each participant’s probability of being a member of each of the three groups, and each participant was classified to the group for which the participant had the highest probability of group membership. Based upon these probabilities, receiver operating characteristic (ROC) curves were created for the classifications for each group, with the goal being to determine how accurately classification-tree models classified all of the participants. The classification tree’s success in classifying participants to each group is illustrated with each group’s ROC curve, which is a plot of sensitivity against the false-positive rate (that is, 1 minus the specificity). The area under the curve (AUC) is an indication of the model’s diagnostic performance. For the target classification (for instance HC), an AUC value is equal to the probability that a randomly chosen HC participant is (correctly) rated or ranked with greater suspicion than a randomly chosen PC or mild AD participant. An area of 0.5 represents chance performance. In building a classification tree model, because of the relatively small sample sizes, k-fold cross validation (with 5 folds) was used. To prevent overfitting the model, the k-fold cross validation stopping rule was used to terminate stepping when improvement in the cross validation RSquare was minimal. The use of classification trees requires no implicit assumption that the underlying relationships between the predictor variables and the dependent variables are linear, follow some specific nonlinear link function (as with generalized linear and nonlinear models) or are even monotonic in nature—that is, they are non-parametric and nonlinear. They are also easy to interpret in that they are based on a series of nested if-then statements.

The qGRE biomarkers in the hippocampus (Dark Matter and Viable Tissue volumes) were used as predictors to build a classification tree, and the results are shown in **Figure 4E** and **Supplemental material (Figures** S2, S3, and S4). Results of a classification tree with the Total Volume as a predictor are shown in Supplementary Figure S5. As indicated in the classification tree (Figure S2, the majority (25 of 34) of HC participants had low Dark Matter volume (less than 704 mm^3^). Of the remaining 9 HCs with Dark Matter volume larger than 704 mm^3^, 8 had high “compensating” Viable Tissue volume (>5782 mm^3^). On the other hand, the majority of participants from the PC group (13 of 18) had Dark Matter volume larger than 704 mm^3^ with 6 having high Viable Tissue volume (>6922 mm^3^), 4 participants with Viable Tissue volume between 5782 mm^3^ and 6922 mm^3^, and 3 below 5782 mm^3^. All 17 participants with mild AD had Dark Matter volume greater than 704 mm^3^ with the majority (9) having low Viable Tissue volume of less than 5044 mm^3^; 5 having Viable Tissue volumes between 5044 mm^3^ and 5782 mm^3^, and only 3 having Viable Tissue volumes larger than 5782 mm^3^ (though less than 6922 mm^3^). Thus, Dark Matter volume was an important biomarker separating participants belonging to HC and PC groups, while Viable Tissue volume played an important role in separating the HC and PC groups from the mild AD group.

ROC-based comparisons of classification results based qGRE metrics versus commonly used tissue atrophy are shown in **Figure 4** and Supplementary Figure S6. Significant differences in the AUC values for the ROC curves were found for HC (p = 0.0304) and mild AD (p = 0.0016). For Dark Matter and Viable Tissue volumes AUC was 0.8676 (0.7662 − 0.9291, 95% confidence interval (CI)) and for Total Volume the AUC value was 0.7273 (0.5989 − 0.8265, 95% CI) with p = 0.0304. The corresponding results for PC were 0.7810 (0.6496 − 0.8729, 95% CI) and 0.7816 (0.6636 − 0.8665, 95% CI), p = 0.9940, and for mild AD, 0. 9519 (0.8838 − 0.8910, 95% CI) and 0.8281 (0.7129 − 0.9033, 95% CI), p = 0.0016.

Data demonstrate that all AUC values for the ROC curves for Dark Matter and Viable Tissue volumes and for Total volumes had p values < 0.0001. For further comparison, for each ROC curve, the Youden Index was used to select the point at which the sensitivity and specificity were optimized. Results using Volumes of Dark Matter and Viable tissue are: for HC, the optimal sensitivity and specificity were 73.53 (55.6 − 87.1, 95% confidence interval (CI)) and 85.71 (69.7 – 95.20, 95% CI), for PC, the optimal sensitivity and specificity were 72.22 (46.5 − 90.3, 95% CI) and 68.63 (54.1 − 80.9, 95% CI), and for mild AD, the optimal sensitivity and specificity values were 100.00 (80.5 − 100.0) and 75.00 (61.1 − 86.0). Difference between these independent ROC curves were assessed using the AUC values their standard errors. The resulting p values were: 0.2143 for HC versus PC, 0.0698 for HC versus mild AD, and 0.0053 for PC versus mild AD.

Corresponding values were calculated for Total Tissue volumes. For HC, the optimal sensitivity and specificity were 76.47 (58.8 − 89.3, 95% CI) and 57.14 (39.4 − 73.7, 95% CI). For PC, the optimal sensitivity and specificity values were 88.89 (65.3 − 98.6, 95% CI) and 62.75 (48.1 − 75.9, 95% CI). For mild AD, the optimal sensitivity and specificity values of 100.00 (80.5 − 100.0, 95% CI) and 50.00 (35.8 − 64.2, 95% CI) were found. Difference between these independent ROC curves were assessed using the AUC values their standard errors. The results were not significant as p values were: 0.4883 for HC versus PC, 0.1841 for HC versus mild AD, and 0.5111 for PC versus mild AD.

The classification results presented in **Figure 4E** can be further improved by including two biological variables (age or gender) in two different classification trees (Figure S3 and Figure S4). By including age, the area under the ROC curve (AUC) increased to 0.8312 for PC but remained practically the same for HC and mild AD. Including gender also increased the area under the ROC curve for PC to 0.8115, but the areas under the ROC curves remained practically the same for the HC and mild AD groups. The confusion matrices show that the combination of the Viable Tissue volume and Dark Matter volume with either age or gender improves identification of participants in the PC group. Our assessments demonstrate that volumes of Hippocampal Dark Matter and Viable Tissue provide useful biomarkers for group classification (HC, PC and mild AD). There are many additional brain areas that could be included in building classification trees, and our expectation is that the inclusion of these additional ROIs will result in improved classification results.

### Association between Dark Matter and direct neuronal count – Preliminary Data

For one participant who expired and underwent postmortem neuropathologic examination, subjective assessment by an experienced neuropathologist found severe loss of neurons in the parasubiculum, the subiculum, and CA1, and modest losses in CA2/CA3 and CA4. Exemplar photomicrographs from CA1 and CA2/CA3 hippocampal subfields are presented in **Figure 5**). These relative neuronal densities align with qGRE measurements that showed a large portion of Dark Matter around the head of the hippocampus (first row in **Figure 5**) – in parasubiculum (25% of Dark Matter), subiculum (18% of Dark Matter), and CA1 (23% of Dark Matter) regions. We found relatively low Dark Matter fraction in CA2/CA3 (3%) and CA4 (10%) regions, which exhibit relative neuronal preservation, as assessed histologically. We also found strong inter-regional association (R^2^=0.78) between in vivo qGRE measurements of Dark Matter fraction and an average neuronal count (N) in 40x objective fields (N=8 in parasubiculum, N=1 in subiculum, N=3 in CA1, N=59 in CA2/CA3, and N=21 in CA4) in the postmortem study. Additional information is presented in Supplemental Table S1.

**Figure 5.**
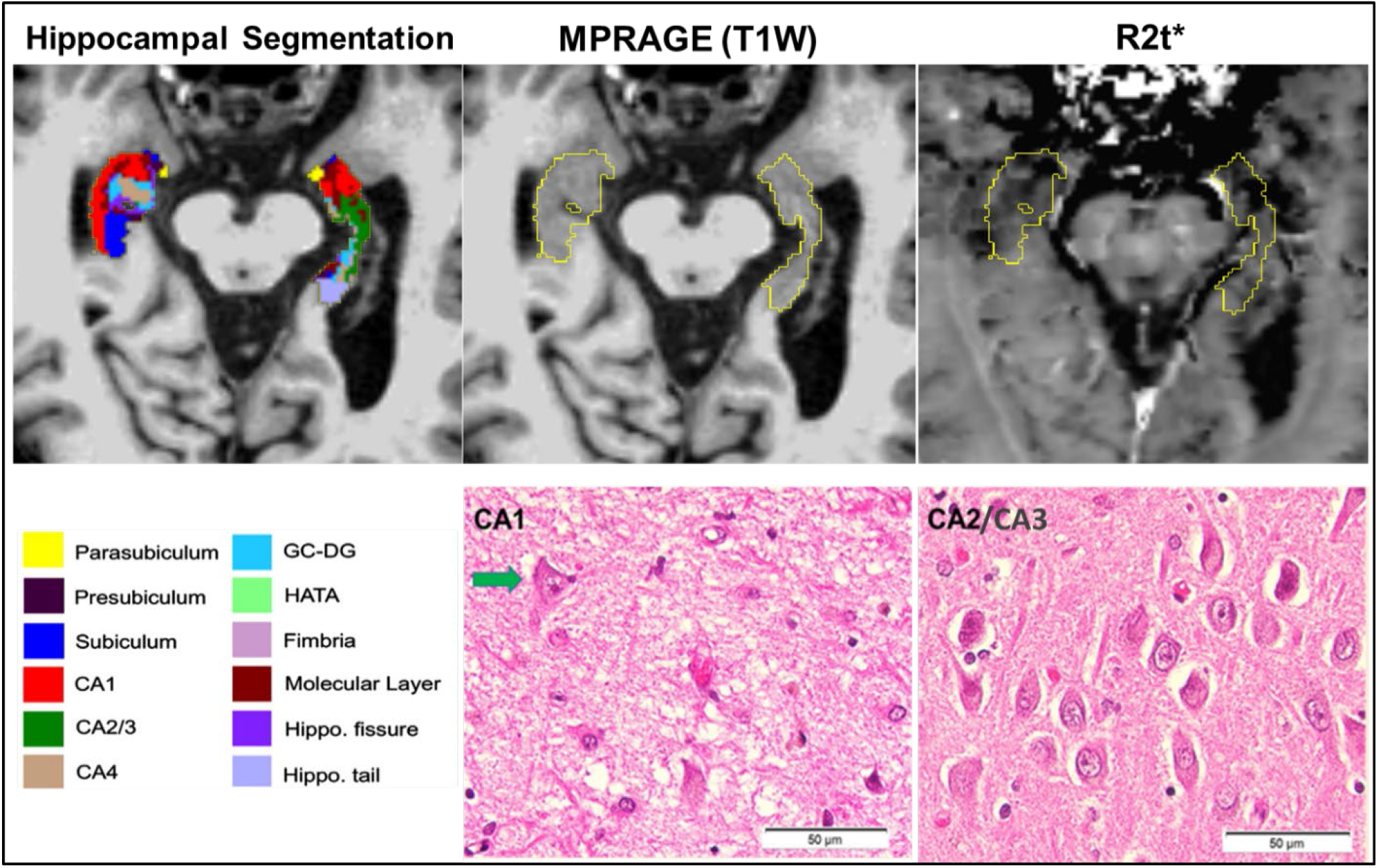
Data obtained from a study participant with a clinical diagnosis of AD dementia (CDR 1). Upper panel – in vivo MPRAGE and qGRE images. Lower panel – postmortem neuropathological examination. qGRE *R2t\** in the hippocampus (outlined in yellow with hippocampal subfields shown in colors, segmented based on FreeSurfer) shows Dark Matter (hypointense lesions with lower *R2t\** values) in parts of subiculum, parasubiculum and CA1, indicating the loss of neurons. This is confirmed by direct neuropathological examination shown in the lower panel obtained from the postmortem studies from this participant. Severe neuronal loss in CA1 (hematoxylin and eosin stain) is reflected by the presence of only one remaining definitively identifiable neuron (indicated by the arrow) within this representative image; relative neuronal preservation is shown in a representative photomicrograph from CA2/CA3. Unlike qGRE *R2t\**, T1-weighted MPRAGE imaging finds the hippocampal region to be practically homogeneous without any obvious intensity contrast. Hence, the data demonstrate a higher sensitivity of qGRE *R2t\** measurements to tissue neuronal loss as compared with standard volumetric measurements. Scale bars are 50 micrometers.

Our preliminary data provide direct validation of the Dark Matter concept, using qGRE to accurately identify brain regions with substantial neuron losses that are not readily detectable on T1-weighted images. This further indicates that in vivo qGRE measurement is sensitive for detecting neuronal loss as compared with T1-weighted based volumetric approach.

## Discussion

This study demonstrates that the qGRE method identifies neuronal loss within hippocampal subfields that is associated with impaired cognition but is not recognized by MRI measurements of tissue atrophy. The onset of neurodegeneration in Alzheimer disease is known to start several years prior to detectable clinical symptoms [2-6] but usually cannot be appreciated, using conventional neuroimaging techniques (i.e. volumetric measurements of tissue atrophy) [6]. A recently proposed A/T/N framework describes a classification system based on the pathological changes accompanying AD progression [24]. With this approach, A refers to amyloidosis, T refers to neurofibrillary tangles, and N refers to neurodegeneration or neuronal injury. Biomarkers for A or T classification include positron emission tomography (PET) and/or cerebrospinal fluid (CSF) measurements, which are well correlated with Braak stages [45]. Losses of neuronal structure or function are defined as neurodegeneration (N) with relevant biomarkers including CSF tau, FDG-PET hypometabolism, and structural MRI (measurement of tissue atrophy). Compared with other techniques, structural MRI (usually T1-weighted MPRAGE) offers high resolution 3D images with good gray/white matter contrast that allow accurate measurement of brain regional volumes and their changes in AD (see recent review [46]). In particular, hippocampal atrophy has been connected to age-related pathology [47] and cognitive impairment [48]; however, as was demonstrated in patients with AD [49], hippocampal regions that have almost isointense contrast on structural MRI might have important variations in measurements of R2t*, reflecting variation in tissue cellular integrity that was correlated with cognitive impairment. This is because T1-weighted imaging only provides the information on global tissue atrophy but is not sensitive to the microstructural changes at the cellular level (i.e. inflammation, loss of neurons, synapses, etc.) in the existing not-atrophied tissue. Consistent with R2t*-based measurement [49], T2-based measurements also identified tissue heterogeneity in hippocampus [50]. Moreover, they found that T2-identified tissue heterogeneity was a better predictor of cognitive decline in MCI participants than mean T2. Several other MRI methods [e.g. diffusion tensor imaging [51], quantitative susceptible mapping [52], arterial spin labeling measurements of the cerebral blood flow [53], Magnetic Resonance Spectroscopy [54]] also reported AD-related changes in brain tissue structure and functioning.

Our approach for evaluation of AD-related tissue cellular damage is based on quantitative Gradient Recalled Echo (qGRE) MRI [31] that allows direct mapping of biomarkers related to human brain cellular composition [32]. qGRE technique is based on a high resolution gradient recalled echo MRI sequence with multiple gradient echoes (available from most MRI manufacturers), the method for separation of cellular specific (R2t*) and BOLD contributions to the R2* decay of the GRE signal [31], and a quantitative relationship between qGRE R2t* metric and tissue neuronal density that was derived in [32] by analyzing an association between maps of R2t* and maps of gene expression profiles obtained from the Allen Human Brain Atlas. This relationship is in concert with a proposed hypothesis [55] that the R2t* parameter can be used as a correlate of cellular structure in healthy brain as well as cellular integrity changes in mild AD [49].

In our study, we used qGRE R2t* mapping to introduce two new quantitative MRI biomarkers to better characterize AD-related, brain-tissue pathology especially at early, preclinical stages of Alzheimer disease – Dark Matter (brain tissue practically devoid of neurons), and Viable Tissue (tissue with a relatively preserved concentration of neurons). The key finding of this study demonstrates that the changes in hippocampal volumes of Viable Tissue and Dark Matter are more sensitive than changes in global hippocampal volume (atrophy) for differentiating healthy control participants from preclinical and mild AD participants, thus providing early biomarkers of neurodegeneration in preclinical AD. Our data also show that the hippocampal volumes of Dark Matter (increased even in preclinical AD stages), and Viable Tissue (decreased with AD progression) are more sensitive correlates of cognitive performance than the global volume of the hippocampus (decreased only in AD stages).

One of the significant consequences of the presence of Dark Matter (and relative preservation of neuronal concentration in the Viable Tissue) is a faster rate of neuronal loss as compared with tissue atrophy. This finding is important due to early observations that the neuron number in layer II of entorhinal cortex determines whether individuals with the neuropathology of Alzheimer disease manifest symptoms: there is little or no neuronal loss in individuals who did not have symptoms of AD during life, whereas individuals with even the earliest symptomatic stages of the disease already had substantial neuronal loss [25, 27]. Hence, it is critical to get in vivo information on neurodegeneration prior to their detection by standard volumetric measurements. Our data demonstrate greater changes in Dark Matter and Viable Tissue (reflecting higher neuronal loss; 31-43% in different hippocampal subfields) as compared with volume loss (10-19% in these subfields) in the mild AD group, relative to HC. This result is consistent with previous findings [25] that reported about 46% of neuronal loss and 29% of volumetric loss (about 1.6x lower than neuronal loss) in CA1 region in the AD postmortem brains compared with brains from those who died with normal cognition.

While no previous report exists on Dark Matter measurements, our findings can be associated with the presence of neurofibrillary tangles (NFT) – a neuropathologic feature most relevant to neuronal loss. In our study, qGRE measurements showed the presence of Dark Matter in the hippocampal tissue even in healthy elderly individuals (age 72±6 years) who are cognitively normal and without amyloid pathology. From NFT perspective, this is consistent with the presence of NFT in cognitively normal older adults [56-58]. Furthermore, the Dark Matter volume increase of about 2.9x in the mild AD group compared with the normal control group is consistent with findings of an autopsy study [59] that reported a ∼2.7x higher concentration of NFTs in the entorhinal cortex of the participants with mild cognitive impairment (MCI) as compared with cognitively normal individuals. Of note, the preclinical designation in our study is not the same as MCI in [59], as our ‘preclinical ‘designation is assigned to participants without cognitive impairment but with amyloid pathology, whereas the term ‘MCI’ classification is based on the presence of cognitive impairment only.

Results shown in **Figure 4** and Supplementary Figure S6 demonstrate advantages of using new qGRE metrics versus commonly used tissue atrophy for participants’ classification between Healthy Control, Preclinical and Mild AD groups. Detailed statistical analyses showed that the AUC values resulting from the ROC curves for Dark Matter with Viable Tissue volumes and for Total volume were all significant (p < 0.0001). However, for Total volume, none of the AUC values were different from one another (p ≥ 0.1841), while for Dark Matter and Viable Tissue volumes, the AUC value for mild AD was significantly higher than that for PC (p = 0.0053) and approached being significantly different from HC (p = 0.0698). In addition, for mild AD, the AUC value for Dark Matter with Viable Tissue volumes was significantly higher than the AUC value for Total volume (p = 0.0016), and the same was true for HC (p = 0.0340). The AUC values for Dark Matter with Viable Tissue volumes and Total volumes were highest for mild AD (respectively, 0.9519 and 0.8281, for which the optimal sensitivities for both methods were 100%; however, the optimal specificity for Total volume (50%) was significantly lower than for Dark Matter with Viable Tissue (75%).

In this study, all hippocampal subfields had lower volume (atrophy) in mild AD when compared with HC group, with the maximum volume loss in the presubiculum, followed by smaller volume reductions in the ML and HP tail regions. These results are consistent with the literature on hippocampal subfields volumetric studies that report lower volumes in different hippocampal subfields in patients with AD compared with cognitively normal individuals [21, 39, 60, 61]. Several imaging [62-64], autopsy [25, 30, 65, 66], and animal [67] studies demonstrate the early involvement of the CA1 region in AD-related neurodegeneration. While our analysis also showed volume reduction in CA1, it did not reach our statistical threshold for our sample of participants, most likely due to insufficient sample size. However, our Viable Tissue volume measurements that consider microstructural neuronal damage, demonstrated differences in the CA1 region between mild AD and HC. In addition, our findings suggest that the volumes of Viable Tissue and Dark Matter can be better candidates than volumetric measurements for predicting cognitive performance. For example, Free and Cued Selective Reminding Test with Immediate Recall (FcSRT) has been widely used to distinguish AD-related dementia from non-AD-related dementia [68, 69], and this neuropsychological test is considered to be the best candidate for episodic memory [70] assessment. A previous study reported a positive but rather weak (r∼0.3) association between hippocampal volume and the FcSRT in a population between 65-80 years [71]. In our participants, the volumetric data also show a positive but weak association between total hippocampal volume and the FcSRT (r∼0.23) but relatively stronger associations with qGRE hippocampal metrics - volume of Viable Tissue (r∼0.37) and Dark Matter fraction (r∼-0.47).

Our study has several limitations. First, the association between R2t* and NDI was derived in [32] for healthy control brains, and this relation might not directly translate for diseased brains. Hence, NDI in AD brain can be treated as an apparent proxy for neuronal density. Other pathological changes in tissue microstructure (e.g. axonal demyelination, depletion of ferritin iron, etc.) that are concomitant with neuronal loss can also potentially lead to reduced R2t*. Further neuropathological studies are required for establishing more detailed information on the relationship between R2t* and tissue cellular composition. In this study, we deduced a NDI only for viable tissue but used a 5.8 s^-1^ threshold for Dark Matter separation from Viable Tissue for all study participants. However, additional analysis presented in Figure S7 shows that our main conclusions are quite stable and the R2t*=5.8 s^-1^ threshold for Dark Matter separation used in our study is also a reasonable criterion for assessment in AD-related participants. Our study had relatively small numbers of participants in each group--a larger study is underway that will further improve the statistical power of our findings. Further, herein we presented a single case of direct histopathological validation of Dark Matter representation as a tissue with significantly lower neuronal content not readily detectable with traditional volumetric measurements. A larger study is also underway to further validate this concept. While in this study we used hippocampal segmentation based on FreeSurfer software, new tools [72] can be more sensitive for specific AD pathology in the hippocampus.

In summary, the key finding of this study is demonstrating that the changes in hippocampal volumes of Viable Tissue and Dark Matter are more sensitive than the changes in global hippocampal Tissue Volume (atrophy) for differentiating healthy control participants from preclinical and mild AD groups, thus providing early biomarkers of preclinical AD pathology. Our data also show that the hippocampal volumes of Dark Matter (increased even in preclinical AD stages) and Viable Tissue (decreased with AD progression) are more sensitive correlates of cognitive performance than the global volume of the hippocampus (decreased only in symptomatic AD stages).

Our approach is based on the qGRE method utilizing a multi-gradient-echo MRI sequence available from most MRI manufacturers and required only about 6 minutes of MRI scanning time. Data analysis can be significantly accelerated with the aid of deep learning method [73], thus opening opportunity for broad research and clinical applications. Combining in vivo qGRE biomarkers of neuronal injury with the current in vivo PET and CSF biomarkers would allow for better understanding and detection of the early pathology in Alzheimer disease and other dementias.

## Methods

### Participants

Seventy (70) participants, ages between 60 and 90 years (73.5±6.6), were recruited through the Knight Alzheimer Disease Research Center (ADRC) and signed the informed consent document. Demographic information available on this cohort is presented in Table 1. Combined MRI and histopathology data from one more additional participant were used for cross-correlation analysis. This study was approved by the Institutional Review Board (IRB) of Washington University School of Medicine.

**Table 1.** Participants’ demographic information and the mean and standard deviation of age and neuropsychometric scores of three groups which are classified based on the CDR and Aβ42 status. ‡ indicates a significant (p<0.05) group difference between HC and mild AD groups, † indicates a significant (p<0.05) group differences between PC and mild AD groups, and + indicates a significant (p<0.05) group differences between HC and PC groups. The Global test is presented as a z-score of combined Animals, Free and Cued Selective Reminding Test (FcSRT), Trail Making A (TMA) and B (TMB), and letter number tests. The rest of the tests are presented as actual individual scores. Note that TMA and TMB are scored such that higher is worse.

| | Total | HC<br>CDR=0 & Aβ<br>status=<br>Negative | PC<br>CDR=0 &<br>Aβ status=<br>Positive | Mild AD<br>CDR=0.5, 1<br>& Aβ status=<br>Positive |
| --- | --- | --- | --- | --- |
| N | 70 | 34 | 19 | 17 |
| Gender<br>(Female/Male) | 31/39 | 19/15 | 8/11 | 4/13 |
| Age | 73.5±6.6 | 72.2±6.1 | 74.7±7.3 | 74.6±6.8 |
| †‡ Global | 0.0±0.8 | 0.4± 0.6 | 0.1±0.8 | -0.8± 0.7 |
| †‡ FcSRT | 27.1±9.2 | 31.9±6.4 | 27.5±8.3 | 17.7±7.7 |
| †TMA | 35.8±15.7 | 30.9±10.4 | 36.2±18.4 | 44.6± 17.9 |
| †‡TMB | 91.2±39.6 | 80.0±30.7 | 85.0±36.4 | 114.8±46.4 |
| †‡Animals | 20.0±6.0 | 21.9±5.7 | 20.2±5.6 | 16.2±5.6 |
| †‡ Letter Number<br>sequencing | 8.9±2.8 | 10.0±2.5 | 9.1±2.3 | 6.8±2.6 |

### Cognitive assessment

All participants of the Knight ADRC complete comprehensive clinical and cognitive assessments. The presence and severity of dementia symptoms was determined using the Clinical Dementia Rating® (CDR®) scale[74, 75]. A CDR score of 0 represents no dementia, and scores of 0.5, 1, 2 and 3 represent very mild, mild, moderate, and severe dementia, respectively, based on exam and collateral source interview. Tests given in the neuropsychological evaluation vary slightly depending on the participant’s age upon entry into the study. Tests administered to all participants include the Free and Cued Selective Reminding Test (FcSRT) [76], Trail Making Part A and B [77], Category Fluency [78] for Animals, and Letter Number Sequencing [79] from the Wechsler Memory Scale-III. A global cognitive score was formed as a z-scored composite of Category Fluency, FcSRT, Trail Making A and B, and letter number sequencing. Episodic memory was represented by the FcSRT. Global cognition test scores were available on 68 participants and the FcSRT scores were available for 62 participants.

### Cerebrospinal fluid (CSF) amyloid measurements

The CSF biomarker Aβ42 test (INNOTEST, Fujirebio, Gent, Belgeium) was performed with a standardized protocol via lumbar puncture. Using the abnormal Aβ42 cutoff value, participants were separated into Aβ positive and negative groups [80].

### Group classification

Seventy participants were classified into three groups based on CDR score and Aβ_42_ status (as shown in Table 1): 1. Healthy control (HC) (CDR=0; Aβ42=negative; n=34); 2. Preclinical AD (PC) (CDR=0; Aβ42=positive; n=19); and 3. mild AD (CDR>0 [CDR=0.5 for 14 participants and CDR = 1 for 3 participants] & Aβ42= positive; n=17). Note that in our consideration we use the term “Preclinical AD” generally accepted in the field as reflecting abnormal biomarkers of amyloid-beta and tau (or the postmortem presence of Alzheimer neuropathology) in asymptomatic persons. This is in part because we and others have demonstrated that preclinical AD is associated with progression to symptomatic Alzheimer disease, whether preclinical Alzheimer disease was detected by amyloid PET [81] or by abnormal CSF concentrations of Aβ42 or tau [82]. This classification corresponds to “Alzheimer’s pathologic change” in the recent NIA-AA framework research criteria [24, 83].

The mean and standard deviation values of cognitive test scores and participants’ ages in HC, PC, and mild AD groups are presented in **Table 1**. There was no difference between groups based on age (p=0.4 between HC and PC, p=0.99 between AD and PC, and p=0.46 between AD and HC). All cognitive tests (except TMA test) exhibit group differences (p<0.05) between HC and mild AD but no group differences between the HC and PC groups (p>0.05). Global cognition, FcSRT, letter number sequencing test..

### Postmortem study

In this study we used data from a participant who was diagnosed with dementia (CDR=1). The participant underwent in vivo qGRE MRI measurements about one year prior to expiration. Neuropathological analysis was performed by a highly experienced board-certified neuropathologist, blinded to any neuroimaging data or formal neuropathologic diagnosis at the time of cell counting. Brain-only autopsy yielded the following neuropathologic findings: hippocampal sclerosis with neocortical and limbic TDP-43 proteinopathy; low Alzheimer disease neuropathologic change (A2, B1, C1 by NIA-AA criteria); mild aging-related tau astrogliopathy (ARTAG); and mild vasculopathy (arteriolosclerosis, cerebral amyloid angiopathy, and atherosclerosis). For this study, neuronal counts were performed on a six-micron-thick coronal section of formalin-fixed, paraffin-embedded tissue from the left hippocampal formation, sampled at the level of the lateral geniculate nucleus, and stained with hematoxylin and eosin (H&E) histochemistry. Three 40x objective fields (each 0.55 mm diameter) selected for counting were evenly spaced but otherwise randomly chosen within each area of interest based on boundaries borrowed from FreeSurfer. Neurons were identified by morphology on H&E stained slides.

### MRI data acquisition

Brain MRI data were acquired at Washington University in Saint Louis. The imaging protocol included the 3D multi gradient-recalled echo sequence and magnetization-prepared rapid gradient-echo imaging (MPRAGE)[84]. Since data were collected over 4 years, four Siemens 3T MRI scanners were used (Siemens, Erlangen, Germany). The Fisher-Freeman-Halton test demonstrated that the three categories of participants (HC, PC, and mild AD) were independent (*P* = 0.2096) of the four different MRI scanners (PET-MR, Prisma, Trio, and VIDA). Details of the statistical analysis are presented in Table S2, therein. Details of qGRE data acquisition are provided in [31]. In brief, sequence parameters were: field of view (FOV) 256×192 mm, resolution 1×1×2 mm^3^ (read, phase, and slab directions), 10 gradient echoes with first echo time 4 ms, echo spacing 4 ms, repetition time TR=50 ms, and flip angle 30°. A phase stabilization echo (navigator) was collected for each line in k-space to correct for image artifacts due to the physiological fluctuations [85]. Standard clinical T1-weighted MPRAGE images (FOV= 256×256 mm, T1/TE/TR=1100/3.37/2000 ms, flip angle=10°, acquisition time= 6 mins, and resolution 1×1×1 mm^3^) were acquired for image segmentation.

### qGRE data analysis

The qGRE MRI method [31, 32] is based on a multi-echo gradient echo sequence theoretical model of GRE signal decay[34, 35] and the relationships between GRE signal decay rate parameters and major elements of brain cellular structure (neurons/neurites and glia cells)[32]. Phase stabilization navigator pulses designed to reduce effects of physiological fluctuations are also incorporated in the sequence[85]. Data analysis was performed with a stand-alone computer with in-house developed programs written in MatLab (MathWorks Inc., Natick, MA, USA). After phase correction, k-space data from each radio frequency (RF) channel were converted to the spatial domain, and the 3D spatial Hanning filter was applied to reduce Gibbs ringing artifacts and signal noise. To achieve optimal model parameter estimations, the multi-channel data (ch = 1, 2, …, M) were combined according to the following algorithm allowing the most accurate model parameters evaluation [86, 87]:

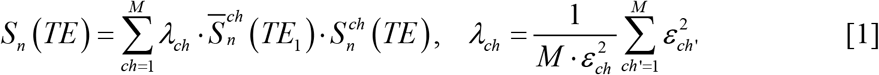

Where 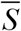 denotes the complex conjugate of S; index n represents the voxel position in space; λ_ch_ are weighting factors, and ε_ch_ are noise amplitudes (r.m.s.).

A theoretical model of BOLD (blood oxygen level dependent) contrast [88] was used to differentiate the contribution of tissue-cellular-specific relaxation (R2t*) and BOLD contributions to the total R2* relaxation [31, 35, 88]:

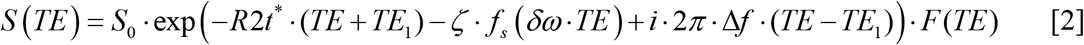

where S_0_ is the signal amplitude; *δω* is the characteristic frequency determined by the susceptibility difference between deoxygenated blood and surrounding tissue; ζ is the volume fraction of deoxygenated blood; nonlinear function *f*_*s*_ (*δω* ·*TE*) accounts for the BOLD effect [88]; and Δf is the local frequency shift, and the function F(TE) describes the effect of macroscopic magnetic field inhomogeneities with respect to gradient echo time TE [89]. Herein, F(TE) was calculated by a voxel spread function (VSF) method [90, 91] using a library-driven approach (45).

### Hippocampal segmentation and co-registration

The hippocampal segmentation was performed on T1-weighted MPRAGE images using Freesurfer6.0 software (Laboratory for Computational Neuroimaging Martinos Center for Biomedical Imaging)[64]. The software provides 12 hippocampal volumes in each hemisphere: parasubiculum, presubiculum, subiculum, cornu ammonis (CA1, CA2/3, and CA4), molecular layer, granule cell layer of the dentate gyrus (GC-DG), hippocampus–amygdala transition area (HATA), fimbria, hippocampal tail (HP tail), and hippocampal fissure. The hippocampal white matter subfields and subfields with lower volumes (HATA, fimbria, hippocampal fissure, and parasubiculum) were excluded from the analysis[21]. The masks of hippocampal subfields in MPRAGE space were registered to the S_0_ qGRE images (that are T1-weighted images) by using FMRIB’s Linear Image Registration tool in FSL software [92, 93]. The resultant transformation matrices were inverted and applied to hippocampal subfield masks of naturally co-registered S_0_ maps of R2t*.

### qGRE metrics generation

The parameters S_0_, R2t*, Δf, ζ, and *δω* in each voxel were estimated by fitting Eq. **[2]** to the complex MR signal in each voxel using a nonlinear least square curve fitting algorithm. The stability of the fitting procedure is described in previous publications [31, 55].

In a recent publication [32], gene expression profiles available from the Allen Human Brain Atlas were used to demonstrate that the network of genes related to neuronal brain structure has an expression profile similar to the R2t* relaxations profile across multiple regions in a human brain cortex. Based on this consideration, the authors deduced a quantitative relationship between the R2t* metric and an index that can serve as a proxy for neuronal density (herein called the Neuronal Density Index, NDI):

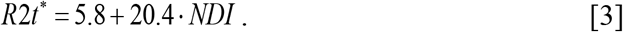

R2t* is measured in sec^-1^, and parameter NDI is dimensionless, which varies from 0 (index for tissue devoid of neurons) to 1 (index for tissue with 100% neurons).

The value of NDI in a healthy adult human brain varies approximately from 0.3-0.7 which correspond to R2t* approximately in the range between 12 s^-1^ and 20 s^-1^; however, data in our study revealed the presence of tissue with R2t* even smaller than 5.8 s^-1^, especially in people with AD. Since NDI > 0, regions of the brain with R2t* smaller than 5.8 s^-1^ represent tissue practically devoid of neurons. Accordingly, with further analysis, we separated brain regions with R2t* smaller than 5.8 s^-1^ from regions with positive NDI (R2t* greater than 5.8 s^-1^). Tissue with R2t* < 5.8 s^-1^ is referred to as “Dark Matter” as it is seen as dark in R2t* images. Accordingly, the volume fraction of Dark Matter in a region of interest (ROI) is characterized by a ratio of Dark Matter volume to the total volume of the selected ROI. The neuron-containing tissue (R2t* > 5.8 s^-1^) is referred to as “Viable Tissue”. Importantly, for all three groups, The R2t* distributions are significantly different between Dark Matter and Viable Tissue as shown in Supplemental Table S3. Note that mean R2t* values in the hippocampal viable tissue are in the range 14.3-15.2 s^-1^ (more than twice bigger than 5.8 s^-1^ threshold) corresponding to NDI index in the range of healthy tissue (as in Figure 1). At the same time, mean R2t* values in the Dark Matter more than twice smaller than 5.8 s^-1^ threshold. The choice of the R2t* threshold (5.8 s^-1^) that separates Dark Matter and Viable Tissue is further justified in Supplemental **Figure S7**. Schematic representation of separating Dark Matter and Viable Tissue is illustrated in **Figure 6** along with examples of hippocampal Viable and Dark Matter Tissue images for three participants belonging to different groups – HC, PC and mild AD.

**Figure 6.**
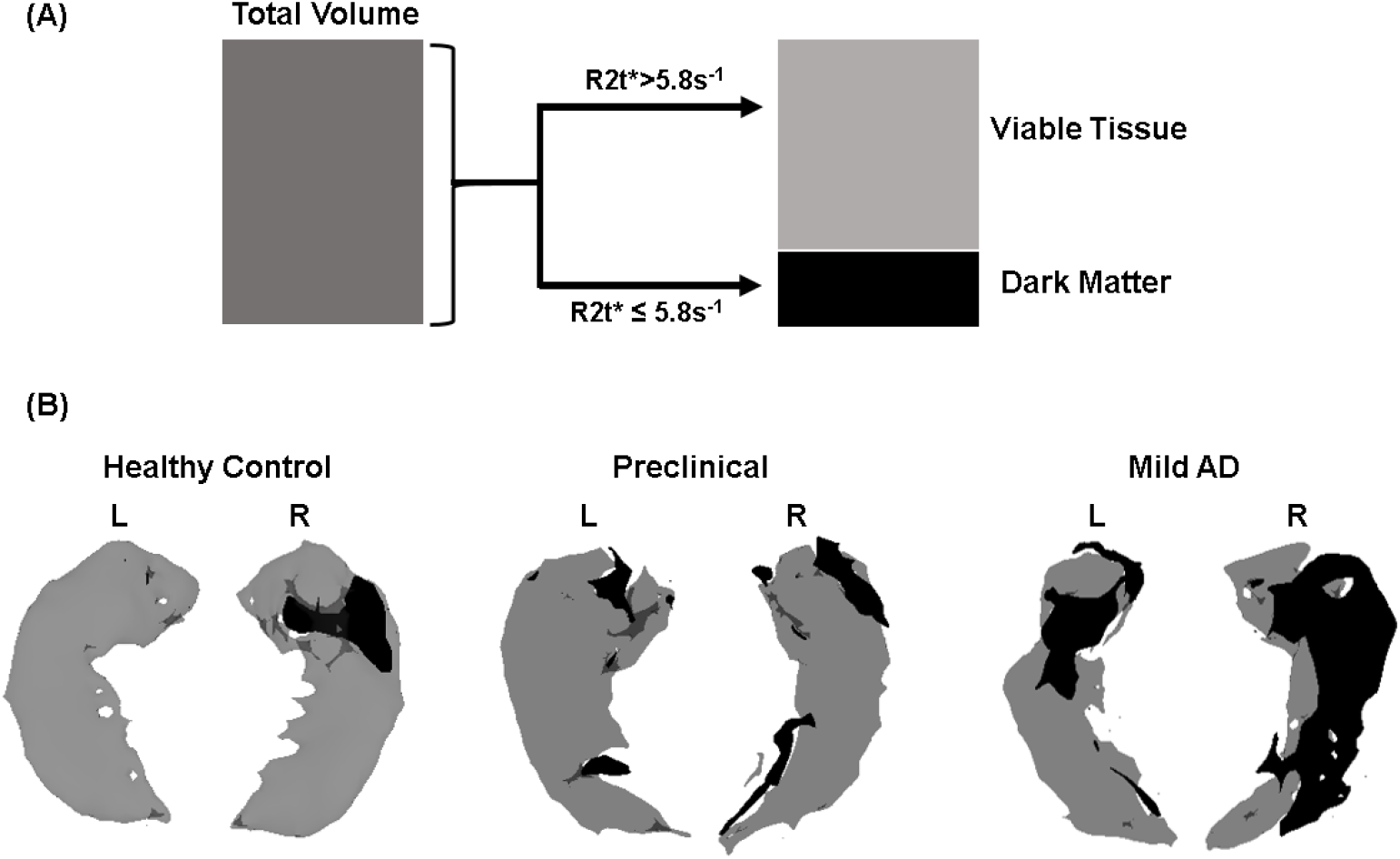
(A) Schematic representation of qGRE biomarkers. Total tissue volume identified on MRI images (e.g. T1-weighted MPRAGE) is separated into two volumes based on qGRE R2t* measurement: Dark Matter - tissue devoid of neurons (R2t*< 5.8 s^-1^) and Viable Tissue – tissue with relatively preserved neurons (R2t*> 5.8 s^-1^). (B) 3D surface views (created by using Slicer 4.5.0 software) of the hippocampus structure of three representative participants from HC, PC, and mild AD groups. Viable Tissue is marked with grey color and Dark Matter is marked with black color.

Note that **Figure 6** shows left/right asymmetry, but this result did not reach significance at the group levels, most likely due to insufficient sample sizes. Hence, in all our consideration, we analyzed data by combining left and right hippocampal subfields.

### Statistical Analyses

For this study, two separate and independent statistical analyses of the collected data were performed. For the first set of analyses, the differences between Viable Tissue and Dark Matter volumes in the individual hippocampal subfields were assessed. For the second set of analyses, global hippocampal measures were used to assess how well the new qGRE metrics (Dark Matter and Viable Tissue volumes) represent clinically important differences between groups as compared with difference based on the hippocampal atrophy (reduction of hippocampal volume).

The first set of statistical analyses was performed using R software (version 22.0, Armonk, NY, United States) and MATLAB (MathWorks Inc., Natick, MA, United States). One-way analysis of variance (ANOVA) with multiple comparisons was performed to assess group differences between HC, PC, and mild AD groups based on qGRE metrics obtained from hippocampal subfields and global cognitive test scores. The associations between qGRE metrics and all cognitive scores were performed using linear regression analysis. From the regression analysis, the correlation coefficient (r) and significance (p) were calculated. Because these analyses were considered preliminary and investigational, a p value less than 0.05 was considered a statistically significant difference and an r value greater than 0.3 was considered a clinically important association between measurements.

For the second set of analyses, differences among groups were assessed with a generalized linear model, and classification tree analyses were used to calculate each participant’s probabilities of being a member of each of the three groups. For the classification-tree analyses, receiver operating characteristic (ROC) curves were created, the areas under the curves (AUCs) were determined for each group and differences between groups determined. In addition, the values for Dark Matter and Viable Tissue volume were assessed for differences from those for Total Volume. For these analyses, the alpha level was set at 0.05. For each ROC curve, it was determined the point at which sensitivity and specificity were maximized. These analyses were performed with JMP Pro Statistical Software Release 15.1.0 (SAS Institute, Inc., Cary, NC) and MedCalc Statistical Software version 19.6 (MedCalc Software Ltd, Ostend, Belgium; https://www.medcalc.org;2020).

## Data Availability

Data associated with this study are present in the paper or the Supplementary Materials. Any additional information can be supplied upon request.

## Acknowledgments

The authors would like to thank Brian Gordon for helpful discussion and Erin Franklin for facilitating and coordinating histopathological evaluations.

## Author contributions

Study design, MRI data analysis: S.K. and D.Y.; MRI data collection: S.K. and T.B.; PET data collection and analysis: T.B.; CSF data collection and analysis: A.F.; Interpretation of cognitive scores: A.A. and J.M.; MRI radiology read: M.G.; Neuropathology study design and analysis: R.P.; statistical analysis S.K., C.H., and D.Y.; manuscript writing: S.K., and D.Y. All authors critically contributed to this work at all stages of project execution and discussion. All authors provided critical comments during manuscript preparation and approved the final version.

## Supplementary Materials

**Figure S1.**
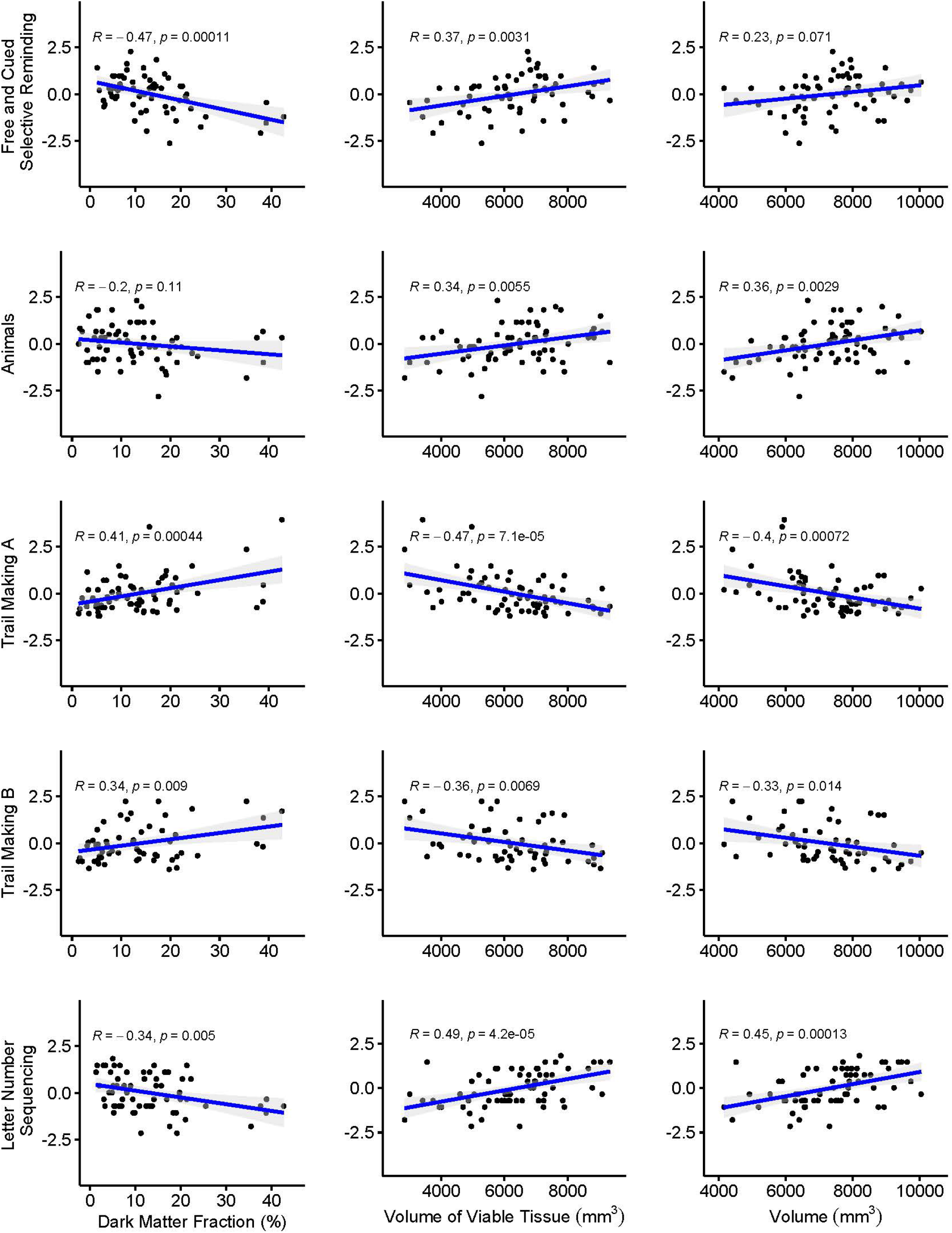
Correlation of individual neurocognitive tests with fraction of Dark Matter, volume of Viable Tissue, and Total Volume of the hippocampus. Each point represents an individual participant.

**Figure S2.**
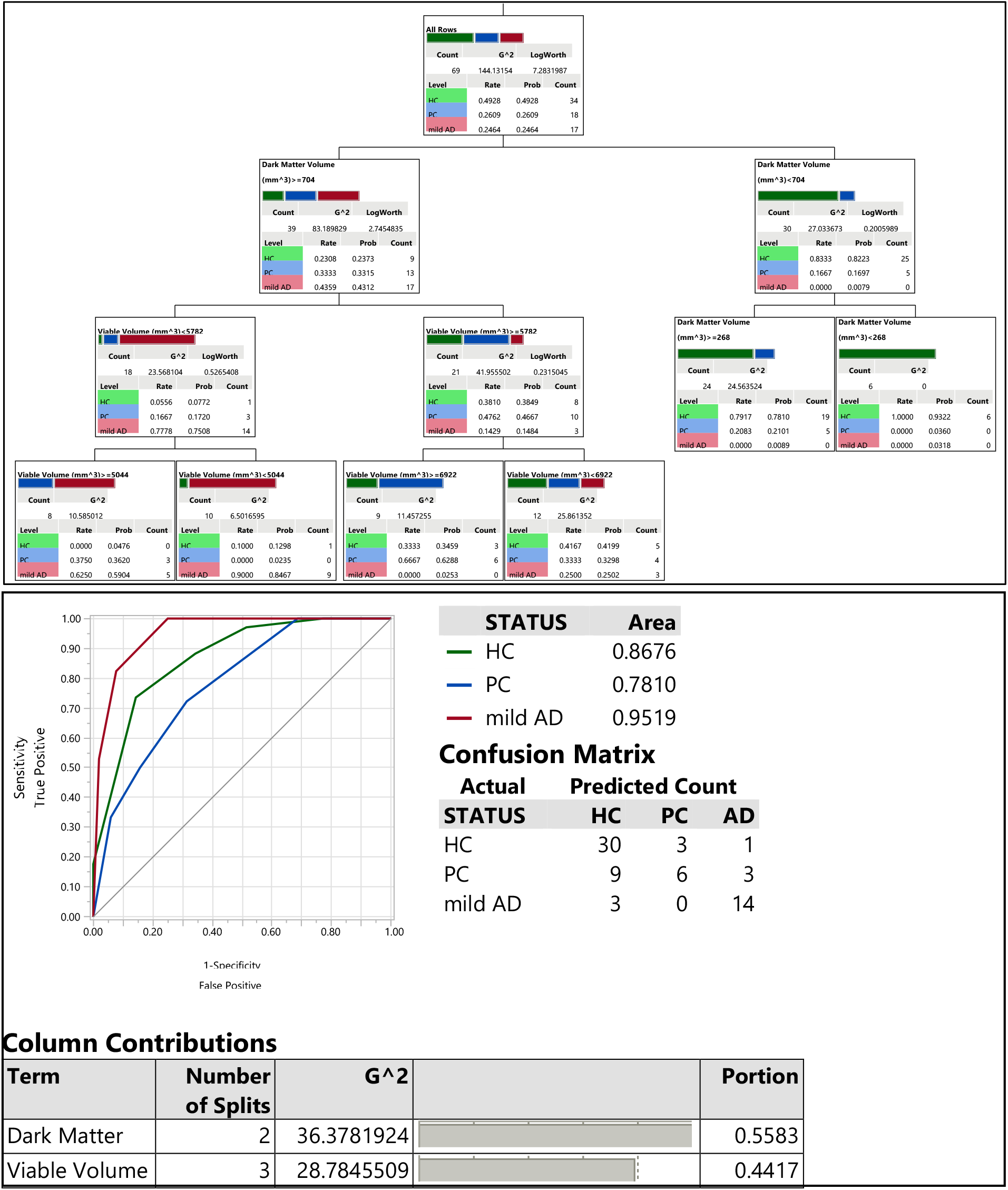
Results of a classification-tree that was produced using global hippocampal Dark Matter volume and Viable Tissue volume variables as predictors. The top panel presents the classification tree diagram with threshold’s, middle panel presents receiver operating characteristic (ROC) curves and areas under the curves (AUCs), a confusion matrix, and bottom panel presents the contribution of variables in the classification of HC, PC, and AD groups. The confusion matrix presents the numbers of correct and incorrect classifications.

**Figure S3.**
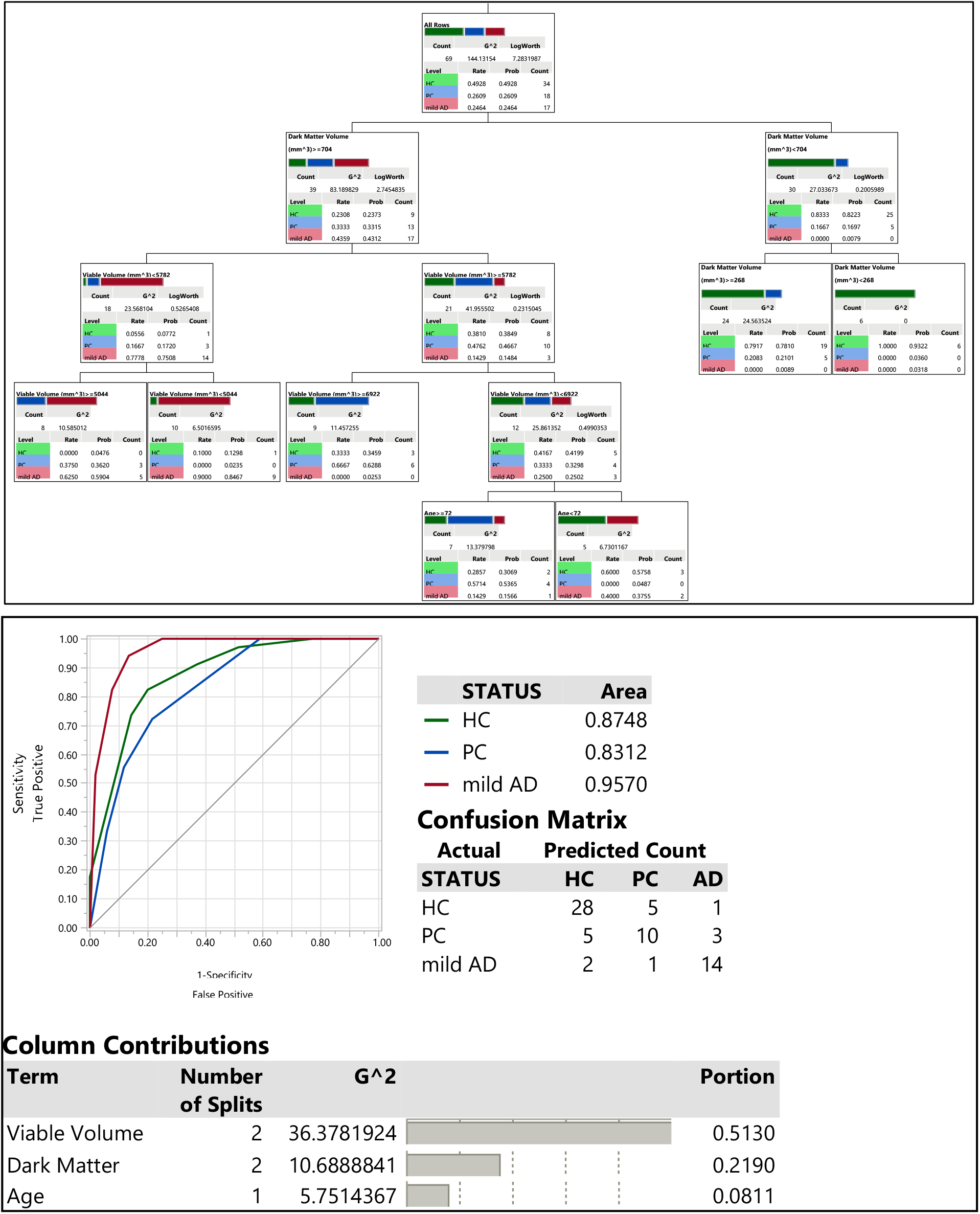
Results of a classification-tree that was produced using global hippocampal Dark Matter volume, Viable Tissue volume, and participant’s age variables as predictors. The top panel presents the classification tree diagram with threshold’s, middle panel presents receiver operating characteristic (ROC) curves and areas under the curves (AUCs), a confusion matrix, and bottom panel presents the contribution of variables in the classification of HC, PC, and AD groups. The confusion matrix presents the numbers of correct and incorrect classifications.

**Figure S4.**
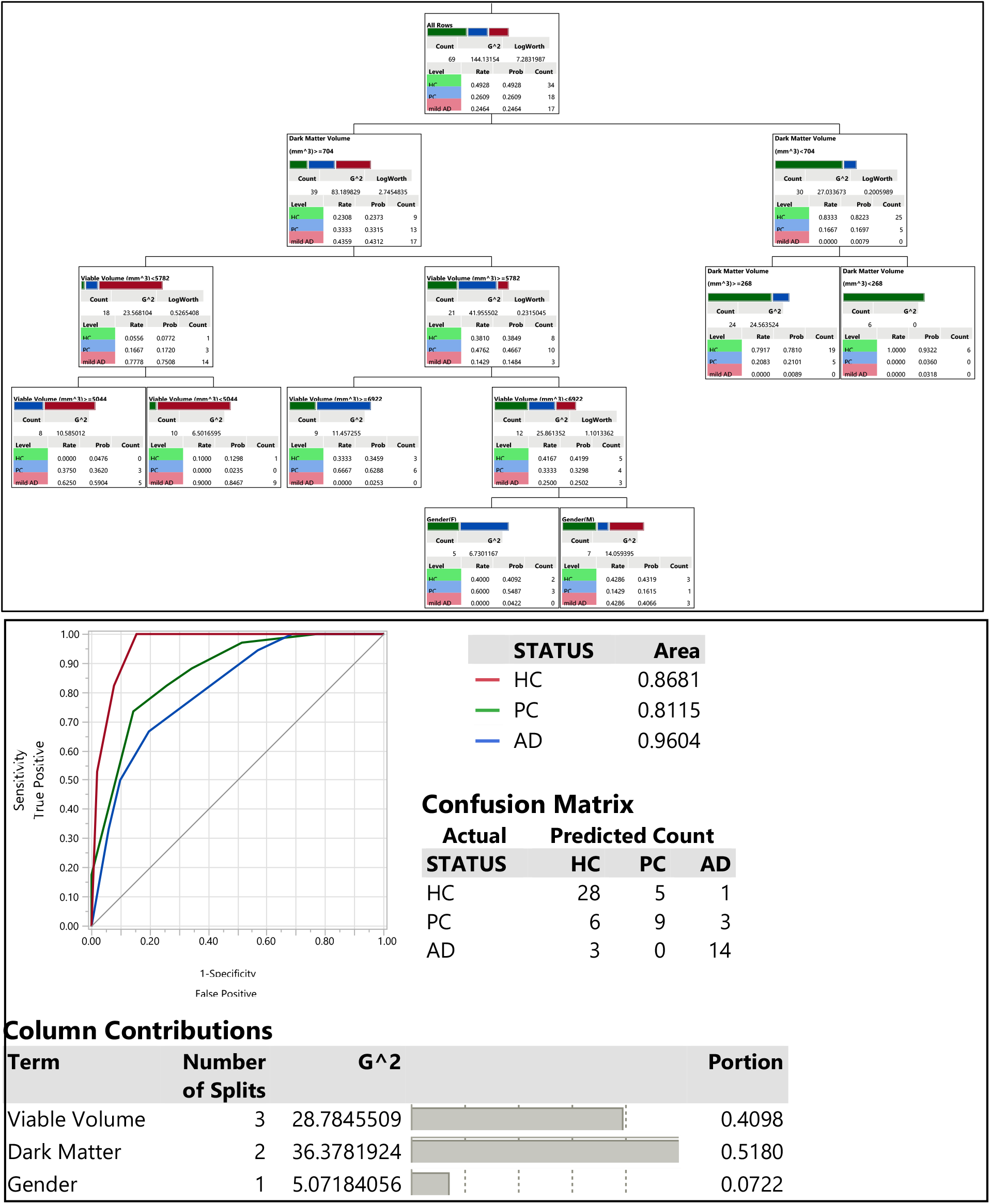
Results of a classification-tree that was produced using global hippocampal Dark Matter volume, Viable Tissue volume, and Gender variables as predictors. The top panel presents the classification tree diagram with threshold’s, middle panel presents receiver operating characteristic (ROC) curves and areas under the curves (AUCs), a confusion matrix, and bottom panel presents the contribution of variables in the classification of HC, PC, and AD groups. The confusion matrix presents the numbers of correct and incorrect classifications.

**Figure S5.**
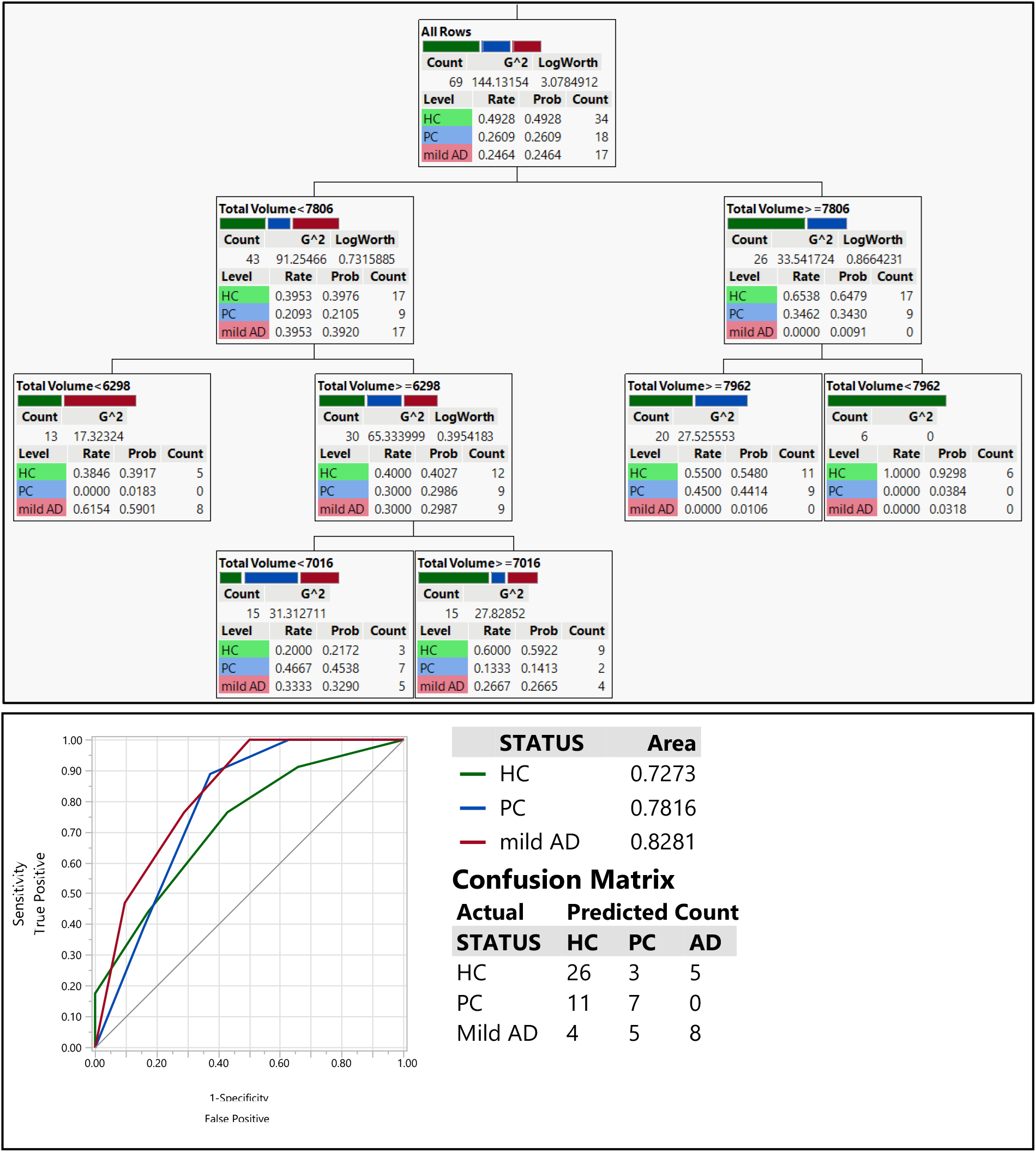
Results of a classification-tree that was produced using hippocampal Total Volume variable as a predictor. The top panel presents the classification tree diagram with threshold’s, lower panel presents receiver operating characteristic (ROC) curves, areas under the curves (AUCs), and a confusion matrix. The confusion matrix presents the numbers of correct and incorrect classifications.

**Figure S6.**
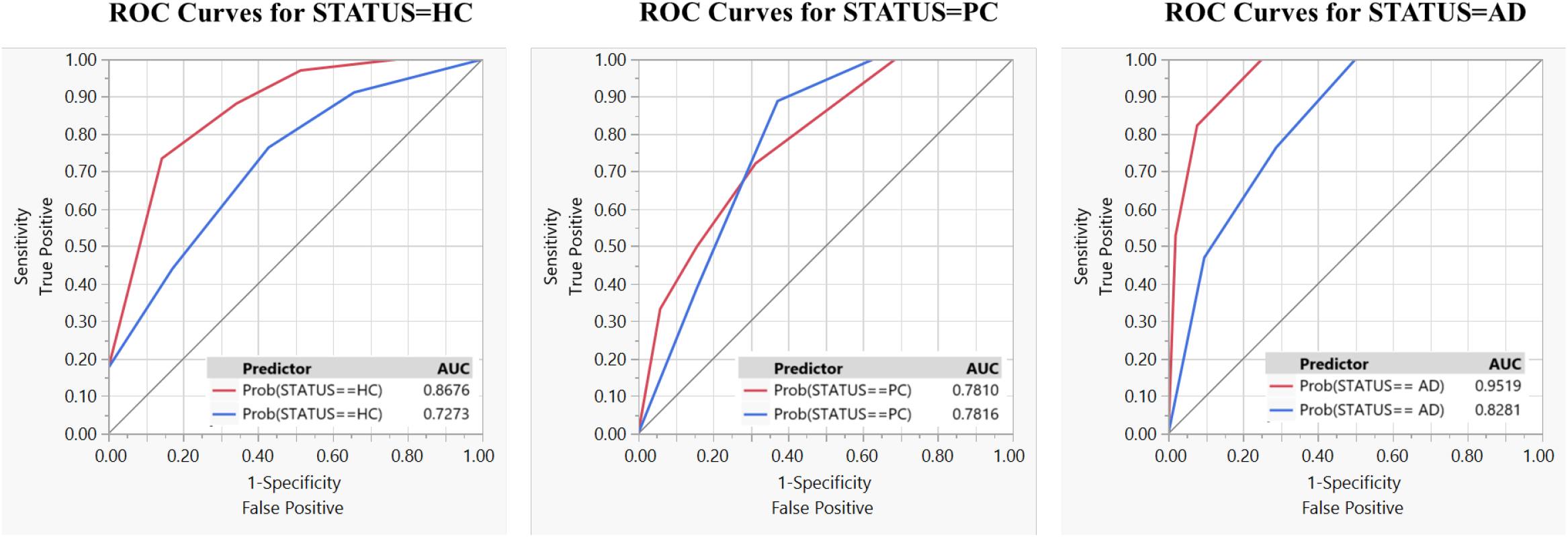
ROC-curve comparisons for classification results based on qGRE metrics versus commonly used tissue atrophy. The comparison of ROC curves created with Dark Matter & Viable Volume versus Total Volume. The red lines represent Volumes of Dark Matter and Viable Tissue used as predictors, and the blue lines represent Total Volume used as a predictor. Insets show corresponding AUC values. Significant differences in the AUC values for the ROC curves were found for HC (p = 0.0304) and mild AD (p = 0.0016). For HC Dark Matter and Viable Tissue volumes AUC was 0.8676 (0.7662 − 0.9291, 95% confidence interval (CI)) and for Total Volume the AUC value was 0.7273 (0.5989 − 0.8265, 95% CI) with p = 0.0304. The corresponding results for PC were 0.7810 (0.6496 − 0.8729, 95% CI) and 0.7816 (0.6636 − 0.8665, 95% CI), p = 0.9940, and for mild AD, 0. 9519 (0.8838 − 0.8910, 95% CI) and 0.8281 (0.7129 − 0.9033, 95% CI), p = 0.0016.

**Figure S7.**
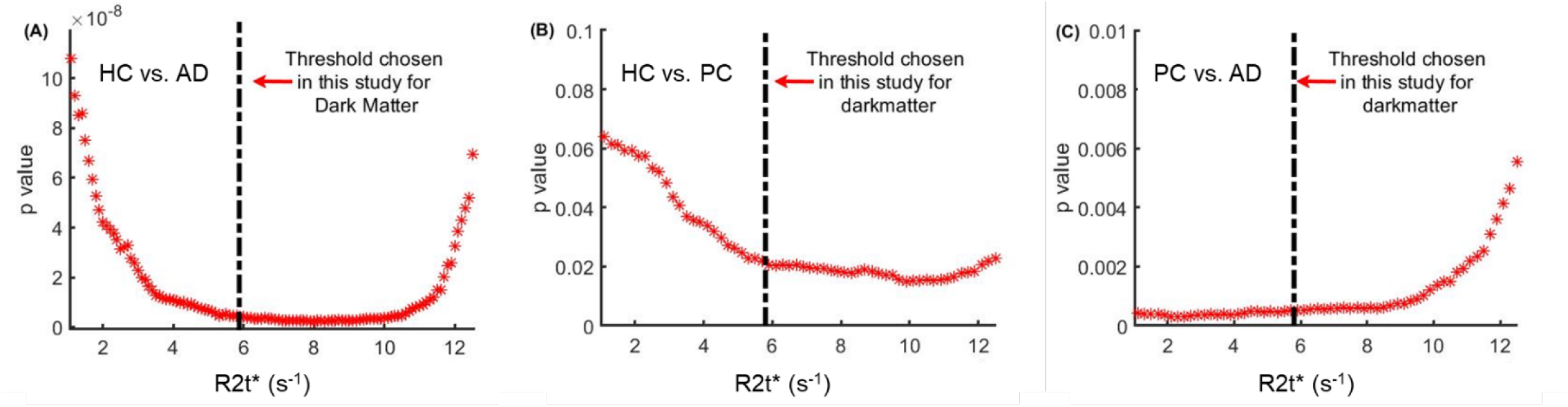
The choice of R2t* threshold for justifying the separation of Dark Matter volume from Viable tissue volume. Plots show dependencies of p value defining significance of group differences between (A) HC and mild AD groups, (B) HC and PC groups, and (C) mild AD and PC groups (using Dark Matter volume as a variable) with varying R2t* threshold (from 1.1 s^-1^ to 12.5 s^-1^ with a step size of 0.1 s^-1^) separating Dark Matter and Viable Tissue. p values obtained from HC and mild AD group comparison, were decreasing with increasing R2t* from 1.1 s^-1^ to 4 s^-1^, remained relatively stable between R2t* of 4 s^-1^ and 10.5 s^-1^ and then started to increase with increasing R2t*. The p values between HC and PC groups were decreasing with increasing R2t* threshold and saturated around 6 s^-1^ till 12.5 s^-1^ and the p values between mild AD and PC groups were small and relatively stable for R2t* values less than ∼9 s^-1^ and then sharply increased above 9 s^-1^. This result illustrates that the R2t*=5.8 s^-1^ threshold for Dark Matter separation is a reasonable criterion for assessment in AD-related participants.

**Table S1.**
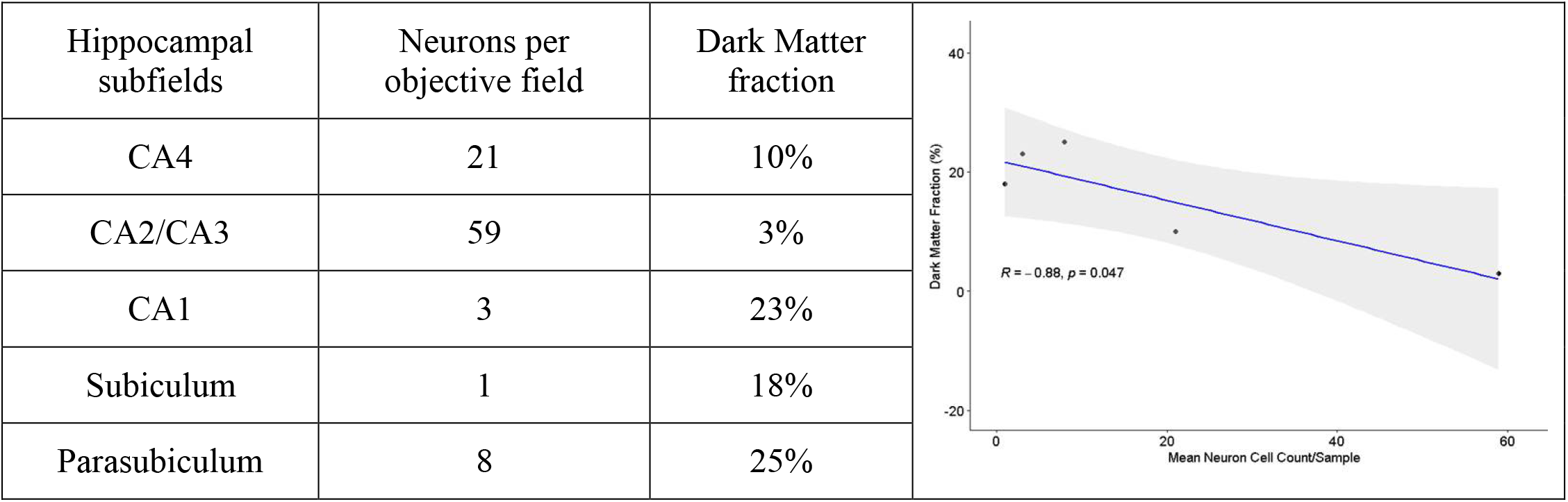
Summary of mean neuronal count and Dark Matter measurements in five hippocampal subfields. Neuron counts were performed by a highly experienced board-certified neuropathologist, blinded to any neuroimaging data or formal neuropathologic diagnosis at the time of cell counting. Three 40x objective fields (each 0.55 mm diameter) selected for counting were evenly spaced but otherwise randomly chosen within each area of interest based on boundaries borrowed from FreeSurfer. Neurons were identified by morphology on H&E stained slides. Plot on the right illustrates an association between actual mean neuronal counts and fractions of Dark Matter in the corresponding regions of hippocampus.

| Hippocampal subfields | Neurons per objective field | Dark Matter fraction |
| --- | --- | --- |
| CA4 | 21 | 10% |
| CA2/CA3 | 59 | 3% |
| CA1 | 3 | 23% |
| Subiculum | 1 | 18% |
| Parasubiculum | 8 | 25% |

**Table S2.** Statistical analysis of the potential influences of MRI scanners on the data obtained in this study. In this study, the data acquired on four different Siemens 3T MRI scanners (PET-MR, Prisma, Trio, and VIDA). The Fisher-Freeman-Hatton test is an extension of the Fisher’s exact test to an unordered r x c table for the three groups of participants (HC, PC, and mild AD) and four different MRI scanners were used (PET-MR, Prisma, Trio, and VIDA). The Count, Total%, Col% (column percent), and Row% correspond to the data within each cell that has row and column headings (such as the cell under PET-MR and HC). The Fisher-Freeman-Hatton test demonstrated that the three groups of participants (HC, PC, and mild AD) were independent (*P* = 0.2096) of the four different MRI scanners (PET-MR, Prisma, Trio, and VIDA).

| Count<br>Total %<br>Col %<br>Row % | PET-MR | Prisma | Trio | VIDA | Total |
| --- | --- | --- | --- | --- | --- |
| HC | 5<br>7.14<br>31.25<br>14.71 | 16<br>22.86<br>64.00<br>47.06 | 4<br>5.71<br>40.00<br>11.76 | 9<br>12.86<br>47.37<br>26.47 | 34<br>48.57 |
| mild AD | 7<br>10.00<br>43.75<br>41.18 | 5<br>7.14<br>20.00<br>29.41 | 1<br>1.43<br>10.00<br>5.88 | 4<br>5.71<br>21.05<br>23.53 | 17<br>24.29 |
| PC | 4<br>5.71<br>25.00<br>21.05 | 4<br>5.71<br>16.00<br>21.05 | 5<br>7.14<br>50.00<br>26.32 | 6<br>8.57<br>31.58<br>31.58 | 19<br>27.14 |
| Total | 16<br>22.86 | 25<br>35.71 | 10<br>14.29 | 19<br>27.14 | 70 |

**Table S3.**
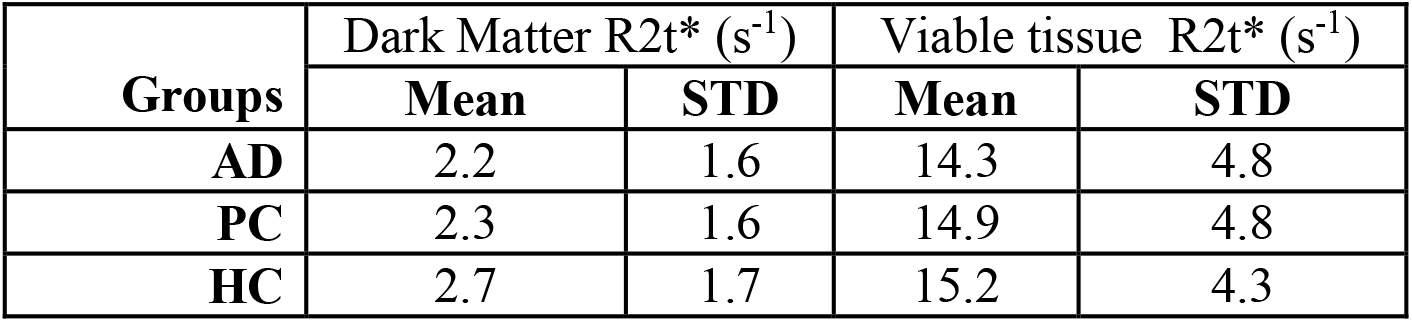
Mean values and standard deviations (STD) of R2t* distributions for three groups in Dark Matter (R2t*<5.8 s^-1^), and Viable Tissue (R2t*>5.8 s^-1^).

